# Polygenic risk of cardiovascular disease manifests in cardiac structure and function

**DOI:** 10.64898/2026.06.07.26354998

**Authors:** Benedetta Felici, Scott C. Ritchie, Saniya Khullar, Carles Foguet, Elodie Persyn, Hasanga D. Manikpurage, Yang Liu, Samuel A. Lambert, Samantha Ip, James H.F. Rudd, Michael Inouye

## Abstract

Cardiovascular diseases (CVDs) are highly heritable, but pathogenesis at the organ and physiological level remains poorly defined. Polygenic risk scores (PRSs), which estimate individual genetic susceptibility to a disease, may allow for the identification of associated subclinical structural abnormalities. Ultimately, identifying where cardiovascular polygenic risk manifests can shape mechanistic hypotheses, guide early interventions, and motivate prevention trials for cardiac remodelling. This study investigated the cardiac features associated with elevated polygenic risk for five common CVDs [heart failure (HF), coronary artery disease (CAD), atrial fibrillation (AF), abdominal aortic aneurysm (AAA) and ischaemic stroke (IS)] using 28 imaging-derived phenotypes (IDPs) from cardiac magnetic resonance imaging of ∼62,000 participants in the UK Biobank cohort. We identified 97 significant PRS–IDP associations as well as 71 IDP–incident disease associations (FDR ≤ 0.05). We further identified 32 significant putative mediators between CVD PRSs and incident disease events, revealing that across CVDs, polygenic risk manifested as distinct patterns in cardiac structures. HF implicated all cardiac chambers, particularly left ventricular and left atrial dysfunction alongside enlarged aorta. AF was characterised by biatrial enlargement and reduced ejection fractions, most prominently in the left atrium. IS and CAD predominantly involved left ventricular hypertrophy, specifically left ventricular wall thickness for IS, whereas AAA was mainly characterised by enlarged descending aorta. Overall, our findings empirically demonstrate that cardiac IDPs partially mediate polygenic risk for CVDs, uncovering a heterogeneous landscape of organ-level mechanisms across CVDs.

## Introduction

Cardiovascular disease (CVD) is the leading cause of mortality globally, accounting for 19.2 million deaths in 2023^1^, with more than 3 million deaths occurring annually in Europe alone^2^. The prediction and prevention of CVDs is therefore a global health priority, particularly for common CVDs such as coronary artery disease (CAD), heart failure (HF), atrial fibrillation (AF), ischaemic stroke (IS) and abdominal aortic aneurysm (AAA).

CVDs comprise heterogeneous pathophysiologies. CAD is marked by the accumulation of atherosclerotic plaque in the intimal layer of coronary arteries, which can lead to heart attacks^3^; while HF arises from dysregulated pumping ability of the heart^4^. AF is characterised by abnormal electrical activity in the atria, leading to impaired atrial contraction and irregular heartbeat^5^. AAA is marked by enlargement of the aorta in the abdomen, defined by an infrarenal diameter greater than 3cm, with risk of aortic rupture increasing with greater diameter^6^. IS is a localised neurological dysfunction due to acute arterial blockage, with risk factors including hypertension and AF^7^. The aetiologies of all these CVDs include substantial genetic components^8–12^.

Polygenic risk scores (PRS) can estimate an individual’s genetic susceptibility to a disease, calculated as a weighted sum of risk alleles across the genome^13^. Across several settings, PRSs have improved risk prediction and stratification^14–19^. For instance, in CAD they can be used to identify individuals with risk profiles comparable to those of familial hypercholesterolemia mutation carriers^20^. While CVD PRSs improve risk assessment, there is a pressing need to identify the pathophysiological pathways through which genetic susceptibility from common variants operates, and to develop targeted therapeutic interventions that disrupt it. Forerunner studies have focused on molecular pathways of polygenic risk using proteomics, e.g. for cardiometabolic disorders^21^, Alzheimer’s disease^22^ and type 2 diabetes (T2D)^23^.

Cardiac structure and function play prominent roles in CVDs, they can be measured by multiple modalities and are targeted by numerous surgical and pharmacological interventions. For example, ventricular stiffness is associated with higher heart failure risk^24^, while reduced descending aortic distensibility and an enlarged left atrium are linked to various cardiovascular conditions, including stroke^25,26^. Such cardiac features are typically assessed using imaging modalities, including magnetic resonance imaging (MRI), cardiac computed tomography (CT), and echocardiography. Recent research has elucidated the genetic basis of cardiac imaging-derived phenotypes (IDPs), highlighting the substantial role genetics plays in shaping organ structure^27–30^. While studies have shown how rare genetic variants for CVDs are reflected in heart structure^31^, including the titin-truncating variants (TTNtv) and eccentric cardiac remodelling^32^, it remains unknown how genetic predisposition to CVDs from common genetic variants manifest in cardiac morphology. Addressing the gap between polygenic susceptibility and organ structure has already advanced our aetiological understanding of schizophrenia and atrial fibrillation^33–35^.

To elucidate whether and to what extent cardiac structure and function mediate PRSs across CVDs, we used cardiac MRI data for ∼62,000 participants in the UK Biobank together with PRSs for five CVDs: HF, CAD, AF, AAA and IS^36–38^. First, to uncover the cardiac features linked to genetic risk for CVDs, we evaluated associations between the CVD PRSs and 28 cardiac IDPs in individuals without prior CVDs. We estimated the total, direct and indirect effects between each PRS and incident events through each IDP, then perform sensitivity analyses to assess the robustness of our results. Finally, we investigated the shared and distinct mediation of PRSs using heart-chamber-stratified analyses. In doing so, we demonstrate that PRSs of CVD operates through cardiac structure and function to varying degrees, which may inform future interventional strategies to reduce CVDs risk in patients with high genetic susceptibility.

## Methods

### Study Cohort

UK Biobank is a cohort of ∼500,000 participants 35–75 years of age, with written informed consent for health-related research, who were recruited through primary care lists between 2006 and 2010^39^. Imaging was conducted at a subsequent follow-up assessment beginning from 2014 onwards, in which 100,000 participants were reinvited and underwent detailed imaging assessments^40^. Ethics approval was obtained from the North West Multi-Center Research Ethics Committee, and participants provided written informed consent for health-related research^39^. This work was conducted under UK Biobank Project 608471.

Starting from the initial cohort of ∼80,000 individuals with available cardiac IDPs, we analysed a subset of 62,516 participants with cardiac IDPs at the first imaging assessment who (1) consented for electronic health record linkage, (2) did not have any prior diagnosis of HF, CAD, AF, AAA and IS and had no prior history of cardiac surgery, (3) had imputed genotype data and were of European genomic ancestry^41^.

### Cardiac imaging traits

Participants underwent cardiovascular magnetic resonance imaging using a standardised protocol^42^. The raw images were subsequently annotated using an automated machine learning pipeline^43^, with the resulting IDPs made available to researchers under UK Biobank category 157. In this study, we utilised set of 28 cardiac IDPs (**Table S1**) obtained from the first imaging assessment timepoint (2014-), retaining only the global measurement for IDPs with multiple segments available. Volume and size based IDPs were indexed to (divided by) body surface area to correct for the well-known correlation between cardiac structure volume and size with body surface area^27,44^. All IDPs were subsequently standardised to improve comparability across traits.

### Cardiovascular disease outcomes

HF, CAD, AF, AAA and IS disease events were defined using a combination of linked electronic health records and self-reported medical history obtained at UK Biobank imaging assessment. Participants were linked to hospital inpatient records (UK Biobank category 2000) and to national death registry records (UK Biobank category 100093) by UK Biobank. Linkage was available from 1997 onwards to 31^st^ March 2023 for hospitals in England, 1981 onwards to 31^st^ August 2021 for hospitals in Scotland, and from 1999 onwards to 31^st^ May 2022 for Hospitals in Wales. Death records and hospital inpatient diagnoses were coded with International Classification of Diseases revision 10 (ICD-10) codes, or ICD-9 codes for events prior to 1996 in Scotland, used to exclude prevalent cases. Surgical procedures were coded with Office of Population Census and Surveys revision 4 (OPCS-4) codes, or OPCS-3 codes for surgeries prior to 1988 in Scotland. Self-reported medical history was collected via a combination of touchscreen questionnaires (UK Biobank showcase category 100044) and verbal interview with a trained nurse (UK Biobank showcase field 20002). Code lists used to define AAA, AF, CAD, HF and IS are provided in **Table S2**.

### Polygenic risk scores

Participants were genotyped using UK BiLEVE arrays and UK Biobank Axiom arrays, and imputed to the UK10K/1000 Genomes and Haplotype Reference Consortium panels, as previously described^45,46^. To prevent confounding by population stratification^47,48^ and maximise power, we restricted analyses to participants whose genotypes clustered with the 1000 Genomes + Human Genome Diversity Panel European reference samples (UK Biobank field 30079)^41^.

PRSs for AAA, AF and HF (**Table S3**) were calculated in UK Biobank from the imputed genotype data using the Polygenic Score (PGS) Catalog Calculator^49^. The PRSs used for AAA and HF were those published by Wang et al. 2023^36^ and were obtained from the PGS Catalog^49,50^, with accessions PGS001784 and PGS001790, respectively. The PRS selected for AF was that of Gunn et al. 2024^37^, with PGS Catalog accession PGS005072. PRSs for IS and CAD were those provided by the UK Biobank (Field ID: 26248 and 26227, respectively)^38^. The PRSs used in the main analyses were selected based on development samples that were independent from the UK Biobank.

For sensitivity analysis, we computed an alternative set of polygenic scores for the same diseases. Since an AF PRS was already available in UK Biobank, we used this score (Field ID: 26212)^38^; for the remaining diseases, we used PRSs obtained from PGS Catalog: PGS002724^18^ for IS, PGS005097^51^ for HF, PGS003973^52^ for AAA, and PGS003725^53^ for CAD **(Table S3)**.

### Study covariates

Several covariates were included in the analysis: sex, age, body mass index (BMI), assessment centre, number of cigarettes smoked per day, self-reported diabetes status, current high blood pressure status, current high cholesterol status, and family history of heart diseases (**Table S1**). High blood pressure status was determined based on participant self-report, systolic and diastolic blood pressure being greater than 140 mmHg and 90 mmHg respectively, self-reported antihypertensive medication usage (touchscreen questionnaire), or use of one of 48 prescription antihypertensives at the imaging assessment (**Table S1**). High cholesterol status was determined based on participant self-report and self-reported lipid-lowering medication usage (touchscreen questionnaire), or use of one of 18 prescription lipid-lowering medications (**Table S1**). Complete-case data were used across all analyses.

### Association between PRSs for cardiovascular diseases and cardiac structure

To investigate the association between PRSs, considered as predictors, and cardiac imaging traits, considered as outcomes, we used robust regression models, to account for the presence of outliers. Specifically, we applied Huber weights using the “MASS” R package^54^ and adjusted the models for age, sex, the first 10 genetic principal components (PCs), assessment centre and genotype array (to account for potential variability in MRI visits and genotype analyses). Given the presence of heteroskedasticity, detected through the Breusch-Pagan (BP) test and visual inspection of residuals, robust standard errors were calculated, using the “sandwich” R package. Across all outcomes, we used the Benjamini-Hochberg (BH) procedure to control for multiple testing, with a false discovery rate (FDR) threshold of 0.05. Two sensitivity analyses were conducted to assess the robustness of results. Firstly, to assess whether the association between PRSs and cardiac structure remained after excluding pathways attributable to established risk factors, we adjusted the analysis for BMI, current high blood pressure, current high cholesterol, daily cigarette consumption, family history of disease and current diabetes. Then, we implemented the same analysis excluding also incident cases, to avoid potential reverse causation. We reported all the results as standardised beta coefficients; namely standard deviation (SD) changes in imaging traits per SD increase in polygenic scores, with corresponding 95% confidence intervals (CIs) and FDR adjusted p-values.

### Association between cardiac structures and incident cardiovascular diseases

Cox proportional hazards models were fitted using the “survival” R package^55^, with the aim to assess the association between cardiac structures and incident cardiovascular disease. Participants were censored due to the end of follow-up, because of death or loss to follow-up^56^. Adjustments were made for polygenic scores for the disease of interest, sex, age, assessment centre, BMI, daily cigarette consumption, current diabetes, current high blood pressure, current high cholesterol and family history of heart disease. The time variable used in the Cox proportional hazards model was calculated as the number of years between the imaging visit and the event of interest. As a sensitivity analysis, we fitted the same models excluding individuals who experienced the event within one year of the imaging visit. This aimed to determine whether the associations between IDPs and incident diseases could be driven by delayed diagnosis, such that some events classified as incidents were prevalent but diagnosed later. Associations were controlled for multiple testing by applying FDR correction across all outcomes jointly. Results are reported as hazard ratio (HR) per SD increase in imaging traits with corresponding 95% CIs and adjusted p-values. Potential violations of the proportional hazards assumption were investigated by inspecting Schoenfeld residuals.

### Counterfactual mediation analysis

To assess the extent that cardiac structure accounted for the association between polygenic risk and incident diseases, we conducted mediation analysis. We used the counterfactual framework to decompose the association between PRSs and incident CVD into components operating through cardiac structure (indirect effects) and components operating through other pathways^57,58^. We sought to make the assumptions of mediation analysis more plausible by conditioning on putative confounders (**Figure S1a**) and by considering BMI, high blood pressure, high cholesterol, and diabetes are post-exposure confounders in sensitivity analyses (**Figure S1b**) (see **Supplementary Methods** for more details). In our setting, post-exposure confounders were variables that affected both cardiac structure and incident CVD, but that might also be influenced by CVD PRSs. In this case, considering them merely as baseline covariates may remove part of the genetic effect we aimed to decompose. However, in the main analyses we targeted conditional associations, considering BMI, high blood pressure, high cholesterol, and diabetes as baseline covariates (**Figure S1a**). This approach was used to isolate the association mediated by cardiac traits, independent of measured risk factors.

We implemented both single- and multiple-mediator analyses using the “cmest” function of the “CMAverse”^59^ package, using 1000 bootstrap resamples. We used the g-formula^60^ approach given its ability to handle post-exposure confounders through randomised analogue effects^61^. Because the outcome model was specified using Cox proportional hazards model, the total natural indirect effect (Rtnie), total natural direct effect (Rtnde), and total effect (Rte) were estimated on the HR scale, whereas the proportion mediated (pm) was estimated on the excess HR scale. As all mediators were continuous IDPs, mediator models were fitted using linear regressions. In primary analysis, we assumed no exposure-mediator interaction (Emint = F); however, this was examined in sensitivity analysis. For continuous exposures, counterfactual mediation analysis is defined with respect to a contrast between two specified exposure values. Consistent with this framework, in the main analysis we set a PRS of 0 as the control value and a PRS of 1 as the treatment value. Sensitivity analyses were then performed using alternative exposure contrasts to assess the robustness of the findings. We used estimation = “imputation” and inference = “bootstrap”. In the main analysis, the following variables were included as baseline covariates (basec), targeting conditional associations: sex, age, BMI, daily cigarette consumption, diabetes, high blood pressure, high cholesterol, family history of heart disease, assessment centre, genotype array, and the first 10 genetic PCs.

For each disease, we considered PRS as exposure, the corresponding incident disease as outcome, and cardiac traits as potential mediators. In single-mediator analysis we included each cardiac trait in turn as a potential mediator. Then, cardiac traits identified as significant mediators in single-mediator analysis (nominal p≤0.05) were subsequently considered jointly in multiple-mediator analysis, to estimate the joint indirect effect mediated by cardiac structure. In the single-mediator analysis, multiple-testing correction was applied to indirect effect tests across all diseases combined, because the indirect effects were the primary quantities of interest. In contrast, for multiple-mediator analysis, we controlled for multiple testing across all tests and all diseases jointly, including direct, indirect, total and proportion mediated effects, because all were of interest and were reported together. Results are reported as HR per SD increase in PRSs with corresponding 95% CIs and FDR-adjusted p-values.

We ran several sensitivity analyses for both single- and multiple-mediator analysis. First, we considered BMI, diabetes, high blood pressure and high cholesterol as post-exposure confounders (postc); while sex, age, assessment centre, genotyping array, family history of cardiovascular disease, daily cigarette consumption and the first 10 genetic PCs as baseline covariates (basec) (**Figure S1b**). Post-exposure confounders were modelled using linear models for continuous variables (BMI) and logistic models for binary variables (the remaining variables). The remaining parameters were identical to those used in the main mediation analysis. However, because post-exposure confounders were present, we estimated randomised analogues of causal effects, see VanderWeele et al. (2014) for more details^61^.

Second, we used alternative polygenic scores to assess whether our findings were robust to the choice of PRSs **(Table S3)**. Third, because causal mediation analyses with a continuous exposure are defined with respect to a specified contrast between two exposure values, we conducted two sensitivity analyses using alternative contrasts. Whereas the main analysis used 0 and 1 as the control and treatment values, respectively, we repeated the analyses by first comparing values from the upper tail of the PRS distribution (3 vs 4) and then values from the lower tail (-4 vs -3). This allowed us to assess whether the mediation results were robust to the choice of exposure values. Then, since in the main analyses we assumed no exposure-mediator interaction, we also reran the analysis including the interaction (Emint = T).

Lastly, to evaluate whether preselecting IDPs based on the single-mediator analysis results influenced the multiple-mediator analysis, we repeated the analysis using both more permissive and more stringent selection criteria. First, we included in multiple-mediator analysis all IDPs, without prior preselection, as potential mediators. Second, we included only IDPs that remained significant in the single-mediator analysis after multiple-testing correction (across 140 tests, FDR≤ 0.05), rather than selecting IDPs based on nominal significance alone.

### Single-mediator analysis adjusting for the other cardiac chambers

Because cardiac IDPs are highly correlated, observed mediation signals may partly reflect correlation with traits from other cardiac chambers, rather than chamber-specific effects. To test this, we estimated indirect effects conditional on measures from the other cardiac chambers (**Figure S1c**).

To capture chamber-level variation, PCs were derived separately for the left ventricle, right ventricle, left atrium, right atrium, and aorta and included in the mediation models as baseline covariates (see **Supplementary Materials** for more details). As a sensitivity analysis, the same analyses were repeated using alternative PRSs for the same diseases.

In the primary analysis, we estimated conditional associations by adjusting for measures of the other cardiac chambers, evaluating whether each cardiac trait mediated the association between genetic risk and incident CVDs independently from other heart chambers. For example, when assessing the mediating role of left atrial ejection fraction, we adjusted for PCs extracted from the left ventricle, right ventricle, right atrium and the aorta (**Figure S1c**). However, given the complex interrelationships among cardiac chambers, simultaneous adjustment for several chambers may introduce overadjustment or collider bias. We therefore adopted a more targeted sensitivity analysis, restricted to chambers with established relevance to each outcome: left ventricle for HF^62–66^ and in CAD^67,68^, and left atrium for AF^35,69^ and IS^70,71^ (**Figure S2**). These established chambers were modelled as both post-exposure confounders and baseline covariates in sensitivity analyses (**Figure S2**). Following the previous example, when assessing the mediating role of left atrial traits in HF, we accounted for the left ventricle as both post-exposure confounder and as baseline covariate (**Figure S2**).

Finally, for HF, given the well-established role of left ventricular ejection fraction^72,73^, we examined whether the mediation signals among left ventricular traits were driven by left ventricular ejection fraction, or whether they persisted even after accounting for it, both as a post-exposure confounder and as a baseline covariate.

## Results

After excluding participants with prevalent CVDs or non-European genetic ancestry, the initial cohort of ∼80,000 individuals with cardiac IDPs was narrowed to a final sample of 62,516 (54% females and 46% males; **Table S4**). At the first imaging visit, the average age was 65.4 years (s.d. 7.8) for females and 66.3 years (s.d. 8) for males, and the average BMI was 26.1 kg/m^2^ (s.d. 4.8) for females and 26.9 kg/m^2^ (s.d. 3.9) for males (**Table S4**). Polygenic scores were calculated for all individuals in the final sample (**Methods**), which included 959 incident cases of CAD, 333 of AF, 310 of HF, 233 of IS and 52 of AAA (**Table S4**). On average, the variables of interest had a missingness rate of approximately 7%. Among the analysed cohort, the average follow-up time was 3.0 years, reaching a maximum of 8.9 years.

A workflow of the study is presented in **Figure 1**. To assess how polygenic risk of CVDs were reflected in cardiac structure and to what extent cardiac imaging traits may account for the association between polygenic risk and clinical events, we conducted four analyses. First, we tested the association between PRSs for five CVDs (HF, CAD, AF, AAA and IS) and 28 cardiac IDPs in the 62,516 UK Biobank participants (**Methods**). Second, we tested the associations between cardiac imaging traits and the five incident CVD outcomes. Third, we applied counterfactual mediation analysis to estimate the extent to which cardiac structures account for the association between polygenic risk and incident CVDs. Finally, we evaluated whether these identified mediation signals could be driven by correlations with other cardiac chambers.

**Figure 1:**
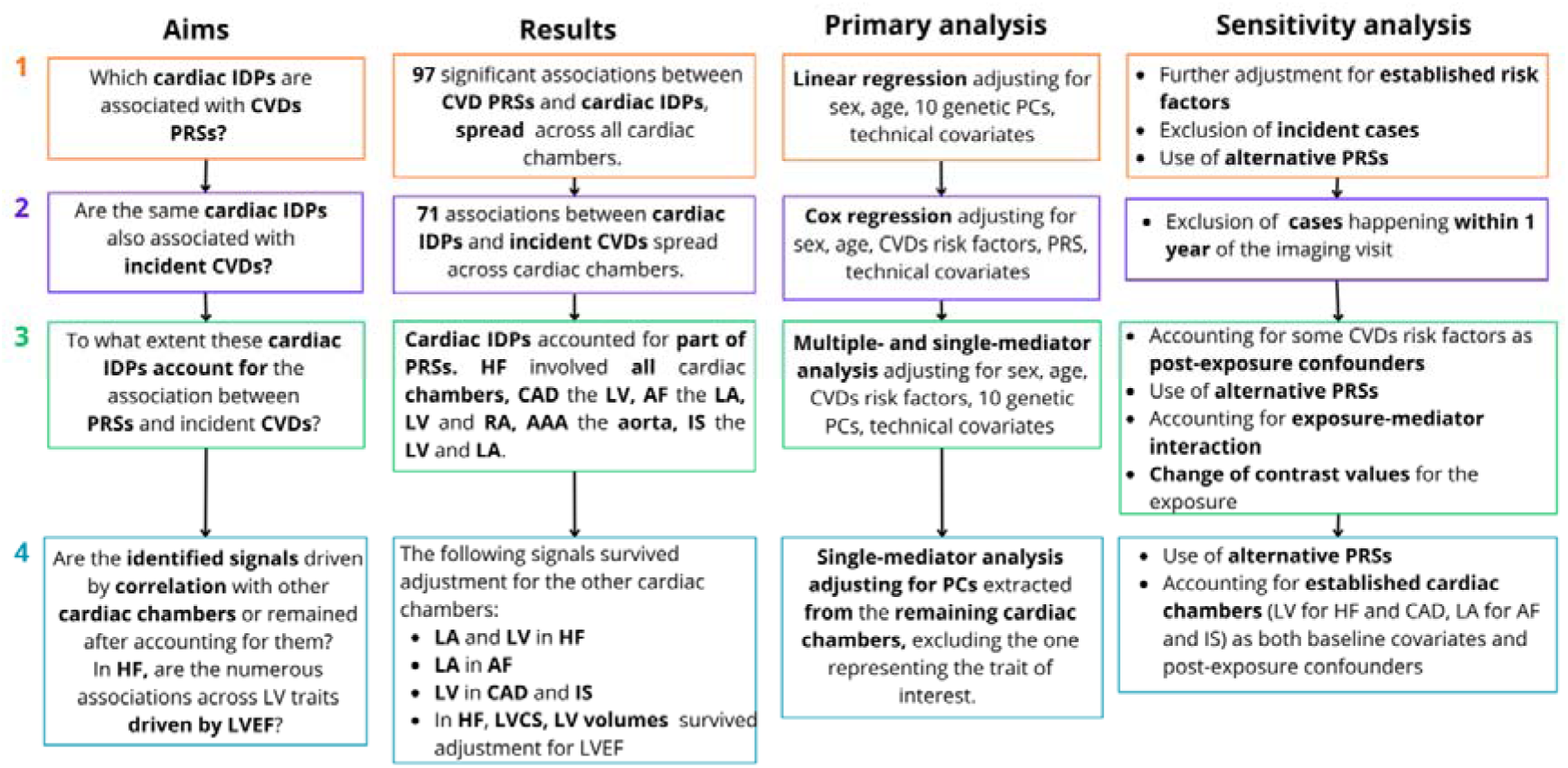
Summary of the study. This figure summarises the aims, the methods (both primary and sensitivity analyses) and the results of our study. **Abbreviations**: LVEF, left ventricular ejection fraction; LV, left ventricle; LA, left atrium; RV, right ventricle; RA, right atrium; LVCS, left ventricular circumferential strain.

In summary, we identified 97 significant associations between the CVD PRSs and cardiac IDPs (across 140 tests, FDR≤ 0.05; **Figure 2a**), 71 between cardiac IDPs and incident diseases (across 140 tests, FDR≤ 0.05; **Figure 2b**). 32 cardiac IDPs emerged as significant mediators between polygenic risk and clinical events in single-mediator analysis (across 140 tests, FDR≤ 0.05; **Figure 3a**). Some cardiac traits demonstrated clear, robust and concordant roles across both polygenic score and corresponding incident diseases. Heart failure was characterised by left ventricular and left atrial dysfunction as well as enlarged aorta; atrial fibrillation by both left and right atrial dysfunction; ischemic stroke as well as coronary artery disease by left ventricular hypertrophy; and abdominal aortic aneurysm by enlarged descending aorta (**Figure 3a**). The indirect effect of polygenic risk acting through cardiac IDPs varied by disease but was substantial when compared with the risk mediated by established cardiovascular risk factors, particularly for heart failure, ischaemic stroke, and atrial fibrillation (**Figure 3b).**

**Figure 2.**
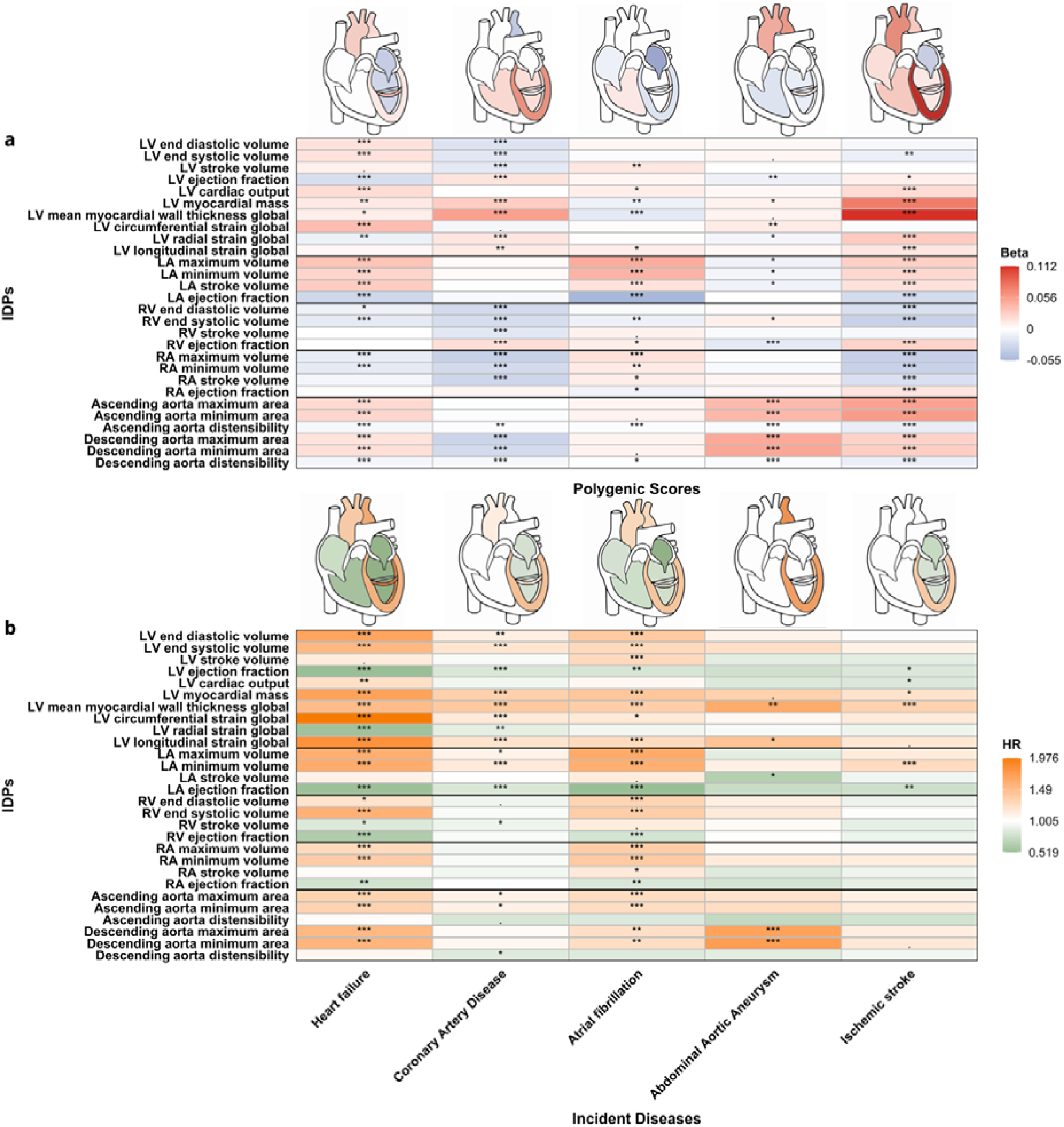
Associations between PRSs, cardiac IDPs and CVDs. In **Panel (a),** the colours represent the beta estimates, namely SD change in imaging traits per SD increase in PRS, derived from robust linear regression adjusted for sex, age, first 10 genetic PCs, assessment centre and genotyping array. PGS Catalog IDs: Abdominal Aortic Aneurysm - PGS001784, Atrial fibrillation-PGS005072, Heart Failure - PGS001790. UKBB Fields IDs: ischaemic stroke (Field 26248), coronary artery disease (Field 26227). In **Panel (b),** the colours represent the HR derived from Cox proportional hazards models adjusting for sex, age, corresponding PRS, daily cigarette consumption, current diabetes, current high blood pressure, current high cholesterol, BMI, family history of diseases and assessment centre. P-values were FDR-adjusted separately within each heatmap: FDR ≤ 0.001 (***), FDR ≤ 0.01 (**), and FDR ≤ 0.05 (*).

**Figure 3.**
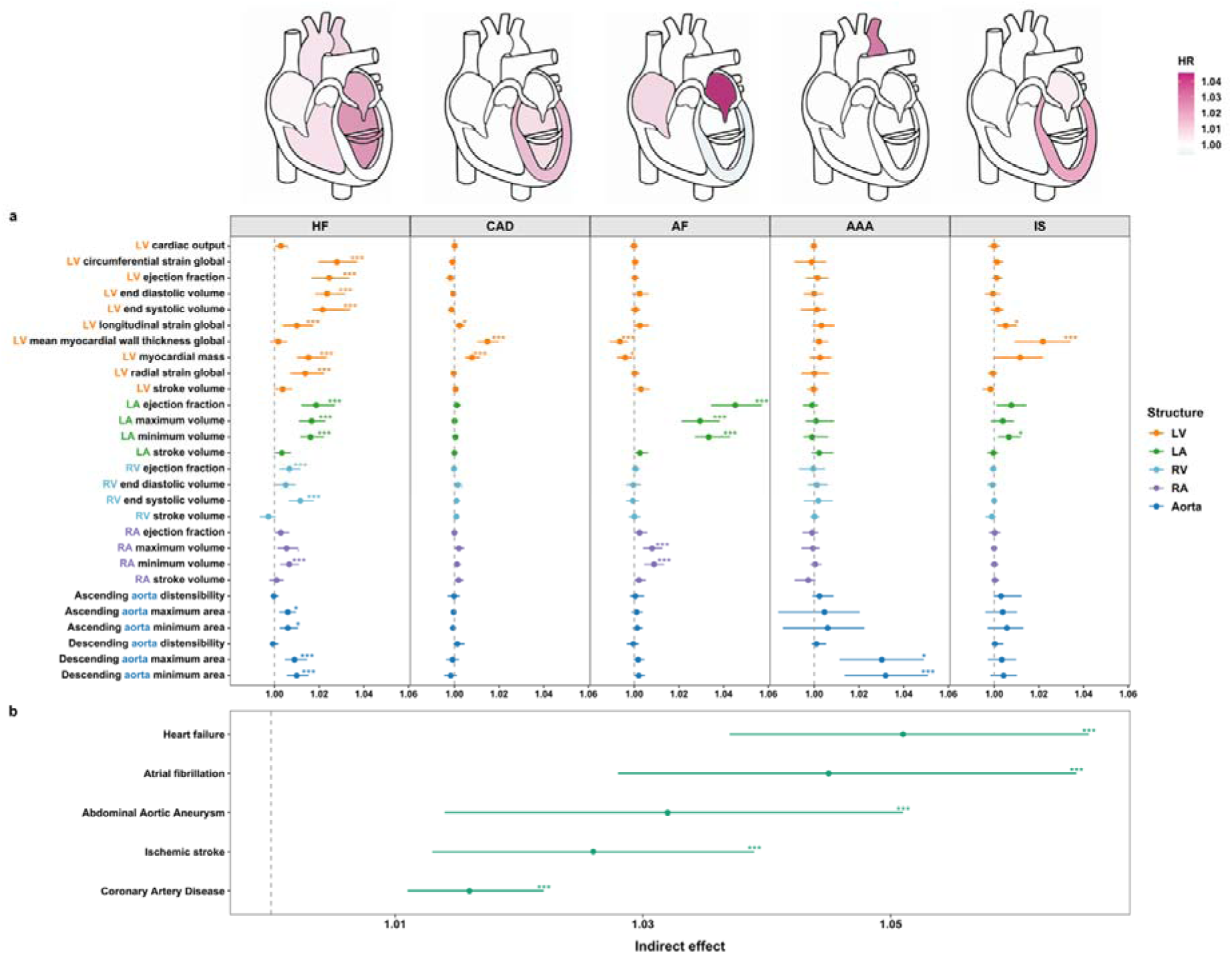
Indirect effects from single- and multiple- mediator analyses. The forest plot in **Panel (a)** represents the indirect effects in HR from counterfactual single-mediator analysis (using Cox proportional hazards models and g-formula in “CMAverse” package). The colours in the heart graphs represent the same HR estimates. For each disease, the exposure was the PRS, the potential mediator was each cardiac trait, and the outcome was the corresponding incident disease. The forest plot in **Panel (b)** represents the indirect effects from multiple-mediator analysis (using Cox proportional hazards models and g-formula in “CMAverse” package). For each disease, the exposure was a PRS, the potential mediators were the cardiac traits identified as significant mediators in single-mediator analysis (p≤ 0.05), and the outcome was the corresponding incident disease. In both panels, the analyses were adjusted for sex, age, the first 10 genetic PCs, assessment centre, genotyping array, daily cigarette consumption, current diabetes, current high blood pressure, current high cholesterol, BMI and family history of disease. CIs were derived using 1000 bootstrap iterations. P-values were FDR-adjusted for multiple comparisons separately within each panel: FDR ≤ 0.001 (***), FDR ≤ 0.01 (**), and FDR ≤ 0.05 (*).

Moreover, mediation analyses accounting for the remaining cardiac chambers suggested that the associations of several cardiac structures with CVD risk were distinct from the other chambers (**Methods**), including left ventricular and left atrial traits in HF, left atrial structures in AF, and left ventricular traits in both CAD and IS (**Figure 4a**). Finally, allowing established CVD risk factors to act downstream of genetic risk (while affecting both IDPs and incident diseases) altered mediation signals: modulating the role of specific cardiac traits while overall increasing the joint indirect effect of cardiac structure across most diseases (**Figure 5**). In parallel, 10.7% of the IDP–disease associations showed discordant estimates, with polygenic risk and incident disease pointing in opposite directions (**Figure 2**). Together, these two findings point to a complex interplay between polygenic scores, CVD risk factors and non-genetic factors in shaping cardiac structure.

**Figure 4.**
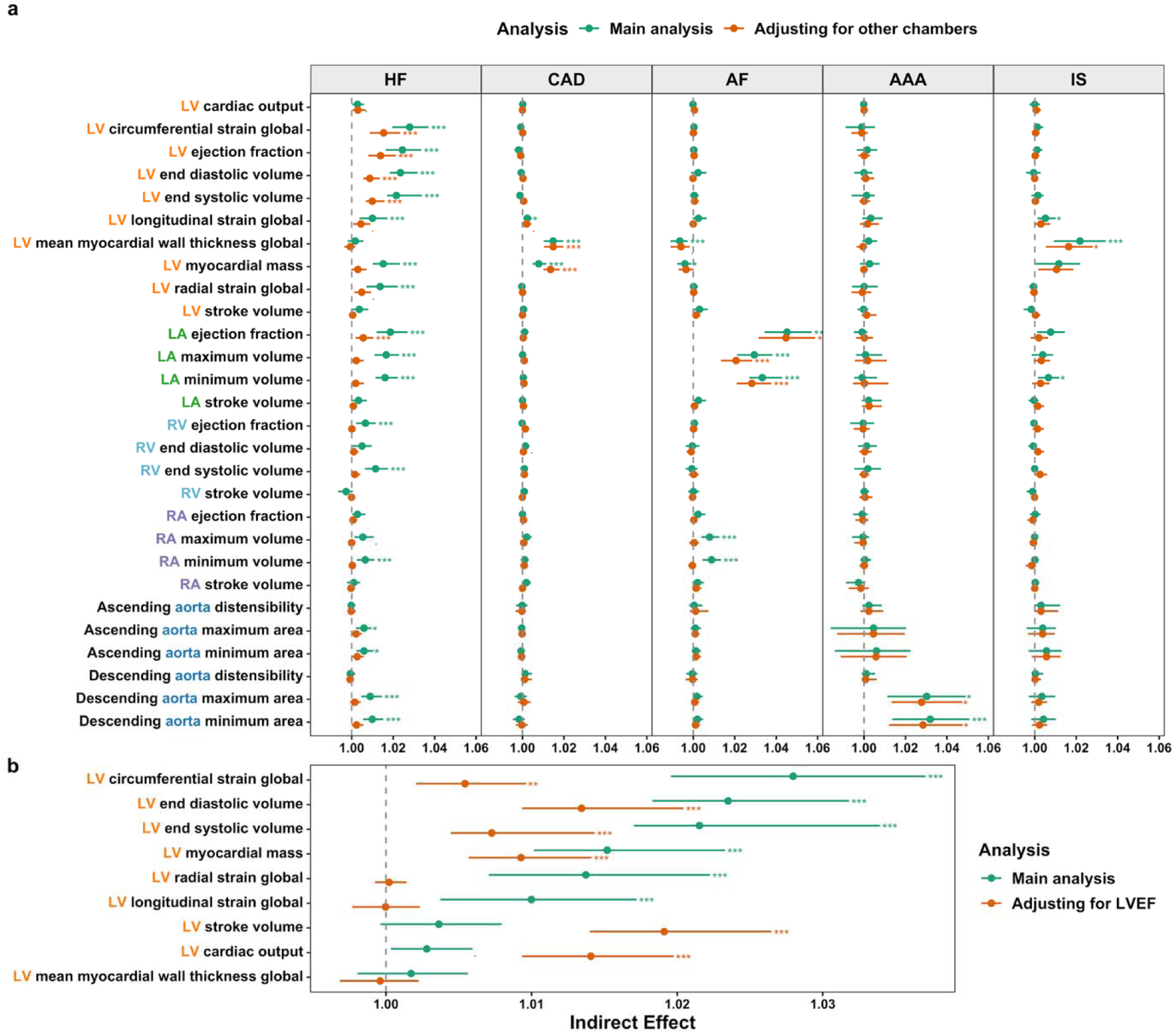
Comparison of primary single-mediator analyses versus models accounting for other cardiac chambers and left ventricular ejection fraction in HF. The forest plot in **Panel (a)** compares the indirect effects from the single-mediator analyses in the main modele versus the secondary model, adjusting for other cardiac chambers. For each disease, the exposure was the corresponding PRS, the potential mediator was each cardiac trait considered individually, and the outcome was the corresponding incident disease. In the main analysis, models were adjusted for sex, age, the first 10 genetic PCs, assessment centre, genotyping array, daily cigarette consumption, current diabetes, current high blood pressure, current high cholesterol, BMI, and family history of disease. In the secondary analysis, models were further adjusted for PCs derived from the other cardiac chambers, excluding the chamber corresponding to the trait of interest. The forest plot in **Panel (b)** shows the indirect effects from the single-mediator analyses of HF and left ventricular traits, comparing the results in the main model versus the model treating left ventricular ejection fraction as a baseline covariate. The exposure was the HF PRS, the potential mediator was each left ventricular trait considered individually, and the outcome was incident HF. In the secondary analyses in this panel, models were additionally adjusted for left ventricular ejection fraction as a baseline covariate. Multiple testing was accounted for within each analysis separately: FDR ≤ 0.001 (***), FDR ≤ 0.01 (**), FDR ≤ 0.05 (*).

**Figure 5.**
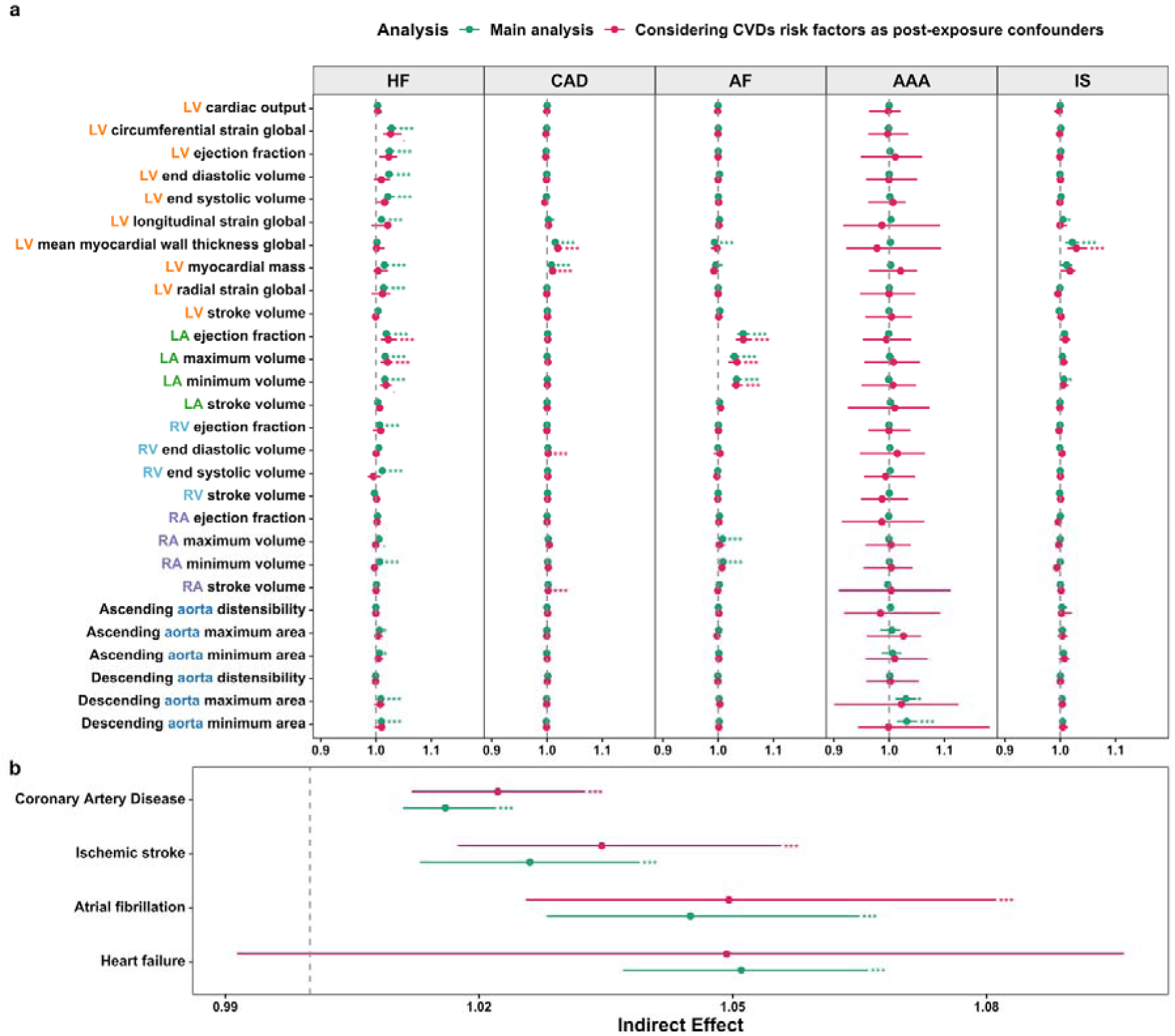
Comparison of primary mediation analysis versus models accounting for CVD risk factors as post-exposure confounders. The forest plot in **Panel (a)** compares the indirect effects from the main single-mediator analyses versus the secondary analysis, accounting for CVD risk factors as post-exposure confounders. For each disease, the exposure was the corresponding PRS, the potential mediator was each cardiac trait considered individually, and the outcome was the corresponding incident disease. In the main analysis, models were adjusted for sex, age, the first 10 genetic PCs, assessment centre, genotyping array, daily cigarette consumption, current diabetes, current high blood pressure, current high cholesterol, BMI, and family history of disease. In the secondary analysis, models were adjusted for sex, age, the first 10 genetic PCs, assessment centre, genotyping array, daily cigarette consumption and family history of disease, while current diabetes, current high blood pressure, current high cholesterol and BMI were modelled as post-exposure confounders. The forest plot in **Panel (b)** compares the indirect effects in the primary multiple-mediator analyses versus the secondary analysis, accounting for CVDs risk factors as post-exposure confounders. The exposure was the PRS, the potential mediators were the cardiac traits identified as significant mediators in single-mediator analysis (p≤ 0.05), and the outcome was the corresponding incident disease. For the multiple-mediator analyses we followed the same approach used in the single-mediator setting, maintaining identical covariate adjustments and treating CVD risk factors as post-exposure confounders in the secondary analyses. Multiple testing was accounted for within each analysis separately: FDR ≤ 0.001 (***), FDR ≤ 0.01 (**), FDR ≤ 0.05 (*).

### Putative mediators of the association between PRSs and cardiovascular disease

#### Heart failure

Heart failure showed widespread significant associations across cardiac structures (**Figure 2**). Of these, 17 partially explained the relationship between polygenic risk and incident HF, spanning ventricular, atrial and aortic measures (**Figure 3a**). From multiple-mediator analysis, the overall indirect effect of the PRS on incident HF through cardiac structure was 1.051 (HR 95% CI: 1.037, 1.066]) (**Figure 3b; Table S5**).

In the left ventricle, the HF PRS was associated with distinctive structural features, characterised by less negative global circumferential strain, lower ejection fraction and decreased global radial strain, as well as greater volumes and mass (**Figure 2a; Table S6**). Impaired left ventricular function, with less negative circumferential strain and decreased radial strain, was also linked to a higher risk of incident heart failure (**Figure 2b; Table S7**). A similar pattern was observed for enlarged volume, mass and reduced ejection fraction (**Figure 2b; Table S7**).

Mediation analysis further indicated that left ventricular strains, volume and ejection fraction accounted for part of the association between the HF PRS and HF onset (HR Indirect Effect: left ventricular ejection fraction 1.024 [95% CI: 1.016, 1.034], left ventricular global circumferential strain: 1.028 [1.020, 1.037]), and left ventricular end diastolic volume: 1.023 [1.018, 1.032]) (**Figure 3a; Table S8**). Which remained statistically significant when accounting for the other cardiac chambers (**Figure 4a; Table S9**). Given the well-established role of left ventricular ejection fraction in heart failure^81,82^, we examined whether signals for left ventricular traits were primarily driven by left ventricular ejection fraction. When accounting for left ventricular ejection fraction both as a post-exposure confounder and as a baseline covariate, the mediation signals for left ventricular volumes and left ventricular global circumferential strain were attenuated but still statistically significant (**Figure 4b; Table S10**).

With respect to left atrial traits, higher HF PRS was associated with left atrial dysfunction, i.e. higher volumes and reduced ejection fraction (**Figure 2a; Table S6).** Left atrial dysfunction was similarly linked with higher risk of HF (**Figure 2b; Table S7**), with left atrial volumes and ejection fraction also partially accounting for the association between the HF PRS and incident HF (HR Indirect Effect: LAVmax: 1.017 [95% CI: 1.011, 1.023], LAVmin: 1.016 [1.011, 1.022], LAEF: 1.019 [1.012, 1.027]) (**Figure 3a; Table S8)**.

After accounting for the other cardiac chambers, the mediating role of left atrial ejection fraction was attenuated by remained significant (**Figure 4a; Table S9**).

The HF PRS was associated with a larger and stiffer aorta (**Figure 2a; Table S6**), and aortic dilation was in turn associated with incident HF (**Figure 2b; Table S7**). Mediation analysis suggested that aortic enlargement partially explained the association between the HF PRS and incident HF (HR Indirect Effect: DAmaxA: 1.009 [95% CI: 1.005, 1.015], DAminA: 1.010 [1.005, 1.015]) (**Figure 3a; Table S8).** However, the mediating role of the aorta showed substantial overlap with that of the other cardiac chambers (**Figure 4a; Table S9**).

#### Coronary Artery Disease

CAD primarily involved the left ventricle, particularly left ventricular hypertrophy, i.e. increased mass and wall thickness (**Figure 3a**). Multiple-mediator analyses suggested an indirect effect of the CAD PRS on the incident disease through cardiac structure of 1.016 (HR 95% CI: 1.011, 1.022]) (**Figure 3b; Table S5**).

Elevated levels of the CAD PRS were associated with greater left ventricular mass and wall thickness (**Figure 2a; Table S6**), which were also linked to a higher risk of CAD (**Figure 2b; Table S7).** Single-mediator analysis suggested that left ventricular hypertrophy accounted for the association between polygenic risk and incident disease (HR Indirect effect, LVM: 1.008 [95% CI: 1.005, 1.012], LVWT: 1.015 [1.010, 1.020]) (**Figure 3a; Table S8)**. Mediation signals for left ventricular hypertrophy remained after accounting for the other cardiac chambers (**Figure 4a; Table S9**).

#### Atrial Fibrillation

AF displayed associations across the left atrium and right atrium, with a particular emphasis on the left atrium (**Figure 3a**). Multiple-mediator analyses suggested an overall indirect effect of cardiac structure on the association between the AF PRS and incident AF of 1.045 (HR 95% CI: 1.028, 1.065) (**Figure 3b; Table S5**).

The AF PRS was associated with larger left atrial volumes and lower ejection fraction (**Figure 2a; Table S6**), which were also associated with a higher risk of AF (**Figure 2b; Table S7**). Single-mediator analysis suggested that the same cardiac traits partially account for the association between polygenic risk and incident disease (HR Indirect effect: LAVmax: 1.029 [95% CI: 1.021, 1.038], LAVmin: 1.033 [95% CI: 1.027, 1.043], LAEF: 1.045 [1.034, 1.057]) (**Figure 3a; Table S8)**. These associations remained robust after adjustment for other cardiac chambers **(Figure 4a; Table S9)**.

The AF PRS was also associated with right atrial dysfunction (**Figure 2a; Table S6**), which was in turn associated with the onset of AF (**Figure 2b; Table S7**). Right atrial volumes partially mediated the association between polygenic risk and incident AF (RAVmax HR Indirect effect: 1.008 [1.004, 1.013], RAVmin HR Indirect effect: 1.009 [1.004, 1.013]) (**Figure 3a; Table S8).** However, right atrial mediation signals overlapped substantially with those of the other cardiac chambers **(Figure 4a; Table S9)**.

#### Abdominal Aortic Aneurysm

Despite small incident case numbers, AAA primarily involved the descending aorta as expected, which showed evidence of an indirect effect in the association between the AAA PRS and incident AAA (HR Indirect Effect: 1.032 [95% CI: 1.014, 1.051]) (**Figure 3; Table S5**).

The AAA PRS was associated with an enlarged and stiffer aorta (**Figure 2a; Table S6**). Larger descending aorta area was significantly associated with increased AAA risk (**Figure 2b; Table S7**). However, this result was not statistically significant after excluding events occurring within one year of imaging visit, potentially due to reduced statistical power (**Table S13**). Single-mediator analysis suggested that the same trait explain part of the association between the AAA PRS and incident AAA (DAmaxA HR: 1.030 [95% CI: 1.011, 1.049], DAminA HR: 1.032 [95% CI: 1.014, 1.051]) **(Figure 3a; Table S8).** Mediation signals were robust to adjustment for the other cardiac chambers **(Figure 4a; Table S9).**

#### Ischaemic stroke

While the brain is the site of injury in IS, thrombotic and embolic causes can depend on cardiac function^74^. We found that IS was mostly associated with the left ventricle, particularly left ventricular wall thickness (**Figure 3a**). Multiple-mediator analysis provided evidence of an indirect effect of the IS PRS on incident disease through cardiac structure (HR Indirect Effect: 1.026 [95% CI: 1.013, 1.039]) (**Figure 3b; Table S5**).

The IS PRS presented 24 associations across all cardiac chambers (**Figure 2a; Table S6)**; it was strongly associated with increased left ventricular wall thickness (**Figure 2a; Table S6**), which was also associated with an elevated risk of incident IS (**Figure 2b; Table S7**). Single-mediator analysis suggested that left ventricular wall thickness partially accounted for the association between PRS and incident IS (HR Indirect effect, LVWT: 1.022 [95% CI: 1.009, 1.034]) (**Figure 3a; Table S8)**, which remained significant after accounting for the other cardiac chambers **(Figure 4a; Table S9)**.

### Cardiac traits showing discordant associations with PRSs and incident disease

A small subset of the IDP–disease associations (10.7%) showed discordant estimates, with polygenic risk and incident disease pointing in opposite directions. For instance, the right atrial and ventricular volumes were negatively associated with HF PRS but positively associated with incident HF (**Tables S6-S7)**. However, controlling for CVD risk factors inverted the direction of the association between HF PRS and right atrial and ventricular volumes, highlighting a complex interplay between genetic susceptibility and established CVD risk factors in shaping heart structure (**Tables S11)**. Similarly, left ventricular wall thickness was inversely associated with the AF PRS but positively associated with incident AF (**Figure 2; Tables S6 -S7**).

Another explanation behind these discordant associations may partly be selection bias, arising from the exclusion of participants with prevalent CVD (**Methods**). Such exclusion may disproportionately remove individuals at high polygenic risk and alter the distribution of polygenic risk and other determinants of cardiac structure in the analytical sample, potentially influencing the observed PRS-IDPs associations.

To further explore the unexpected inverse association between AF PRS and left ventricular wall thickness, which was also observed in single-mediator analysis, we examined both genetic and non-genetic determinants of left ventricular wall thickness. We found that the Townsend Deprivation Index and AF PRS were associated with left ventricular wall thickness in opposite directions, supporting the possibility that geneticand non-genetic factors may influence left ventricular wall thickness, and potentially AF risk, in different ways (**Table S12**).

### Considering CVD risk factors as post-exposure confounders of the association between cardiac structure and CVD

We repeated the mediation analyses treating BMI, high blood pressure, high cholesterol and diabetes as post-exposure confounders, allowing them to lie downstream of genetic risk while confounding the association between cardiac traits and diseases (**Methods**). Loss of statistical significance after this adjustment may partly reflect reduced statistical power and precision arising from the greater model complexity, rather than necessarily indicating absence of mediation.

For HF, treating CVD risk factors as post-exposure confounders attenuated the mediation signals from left ventricular and aortic traits, while strengthening the signals from left atrial ejection fraction and left atrial maximum volume (**Figure 5a**, **Table S13**). In the multiple-mediator model, the indirect effect was similar in magnitude but no longer statistically significant, suggesting reduced precision rather than a substantial change in the estimated effect (**Figure 5b**, **Table S14**).

Turning to CAD, after accounting for CVD risk factors as post-exposure confounders, additional right-sided cardiac traits emerged as potential mediators, including right ventricular end-diastolic volume and right atrial stroke volume, alongside the already identified left ventricular mass and wall thickness (**Figure 5a**, **Table S13**). The overall indirect effect in the multiple-mediator model increased slightly (**Figure 5b**, **Table S14**). For AF, mediation signals involving the right atrium were attenuated, whereas left atrial ejection fraction as well as left atrial maximum volume remained significant putative mediators (**Figure 5a**, **Table S13**). The overall indirect effect from cardiac structure also rose marginally in the multiple-mediator model (**Figure 5b**, **Table S14**).

In the case of AAA, no mediation effects remained statistically significant after accounting for CVD risk factors (**Figure 5a**, **Table S13**). This may partly reflect reduced statistical power due to the greater model complexity and the relatively small number of AAA events. In IS, both the indirect effect from left ventricular wall thickness and the overall indirect effect from the multiple-mediator model increased modestly after accounting for post-exposure confounding (**Figure 5**, **Table S13-S14**).

### Sensitivity analyses

Associations between PRSs and cardiac IDPs were broadly consistent after excluding incident cases, adjusting for established CVD risk factors, and using alternative PRSs for the same diseases (**Supplementary Figure S3**, **Table S11, S15-S16**). Associations between cardiac traits and incident disease were also generally unchanged after excluding events occurring within one year of imaging; however, CIs widened, leading to reduced statistical power for certain associations, particularly for AAA and IS (**Supplementary Figure S4, Table S17**). For instance, the association between left ventricular wall thickness and incident IS decreased slightly from 1.33 (HR 95% CI: 1.16, 1.53) to 1.18 (HR 95% CI: 1.00, 1.40) losing significance (**Supplementary Figure S4, Table S17**).

Mediation estimates were similarly consistent across a range of sensitivity analyses, including using alternative PRSs, varying the treatment and control values across the upper and lower tails of the PRS distribution, and allowing for exposure-mediator interaction. In single-mediator analysis, estimates of indirect effects were generally robust, although statistical significance was attenuated in some more complex models, particularly when post-exposure confounders were included (**Supplementary Figure S5, Tables S18-S21**). In multiple-mediator analysis, indirect effects remained broadly consistent across all sensitivity models, including variations in selection of mediators (**Methods**, **Supplementary Figure S6, Table S14**). While statistical significance attenuated in more complex models, particularly when post-exposure confounders were included, point estimates changed minimally (**Supplementary Figure S6, Table S14**). For HF, the indirect effect decreased from 1.051 (HR 95% CI: 1.037,1.066) in main analysis, to 1.049 (HR 95% CI: 0.991,1.096) when accounting for post-exposure confounding (**Supplementary Figure S6, Table S14**).

## Discussion

In this study, we sought to elucidate how CVD PRSs manifests in differences in cardiac structure and function. To this end, we analysed associations between CVD PRSs, cardiac IDPs and incident CVD events using linear regression, Cox proportional hazards models and counterfactual mediation analysis. We identified 97 significant associations between CVD PRSs and cardiac IDPs, 71 associations between cardiac IDPs and incident CVDs, and 32 cardiac traits resulted as significant mediators between PRSs and incident CVD events. Several mediation signals persisted after accounting for the remaining cardiac chambers, suggesting partially distinct contributions.

Overall, multiple-mediator analyses suggested a substantial indirect effect of PRSs through cardiac IDPs, though the magnitude of the indirect effect varied from a HR of 1.051 (for HF) to 1.016 (for CAD). This heterogeneity may reflect differences in diseases pathophysiology as well as variation in the performance of the selected PRSs.

For comparison, a mediation analysis considering total cholesterol, high-density lipoprotein cholesterol (HDL-C), BMI, systolic blood pressure and diabetes as mediators of the association between CVD PRS and cardiovascular disease reported a combined indirect HR of 1.03, with HDL-C alone showing an indirect HR of 1.02^75,76^. Similar indirect effects have been reported for apoB, apoA I and blood pressure as mediators of the association between coronary heart disease (CHD) PRS and CHD risk^76^.

Polygenic risk of HF manifested in all cardiac chambers, with left ventricular, left atrial and aortic traits emerging as putative mediators. Associations involving left ventricular traits and left atrial ejection fraction remained robust after adjustment for the other cardiac chambers. The well-established role of the left ventricle in HF is supported by a recent Mendelian randomization study and is consistent with prior studies showing genetic correlation, colocalization, and pathway enrichment between HF and left ventricular traits^62,63,65,66,77^. Importantly, left ventricular circumferential strain and volume remained robust after adjustment for left ventricular ejection fraction, indicating that the contribution of the left ventricle cannot be reduced to ejection fraction alone^78–82^.

HF additionally showed associations with left atrial volume and ejection fraction. This supports an important role for the left atrium in HF, and it is consistent with prior evidence from colocalization, from analyses using PRS for the left atrium, and studies of heart failure subtypes^35,65,83,84^. Interestingly, left atrial traits demonstrated greater robustness than left ventricular ones when accounting for CVD risk factors as post-exposure confounders, suggesting that chamber-specific remodelling may be driven by distinct mechanisms. This aligns with recent findings in heart failure with reduced ejection fraction (HFrEF), where proteomic data similarly suggested distinct pathways underlying atrial and ventricular dysfunction^85^.

Surprisingly, aortic traits emerged as putative mediators of HF polygenic risk, which may reflect a shared genetic susceptibility to aortic remodelling and impaired “windkessel” function^26^. Previous studies have shown that larger aortic diameter was associated with increased risk of cardiovascular mortality, while echocardiographic studies have linked aortic root remodelling to the risk of incident HF.^86,87^

CAD was consistently associated with left ventricular hypertrophy, i.e. increased mass and wall thickness. Prior studies have linked increased left ventricular mass to CAD and related cardiovascular outcomes^67,68^. Although hypertension may partly explain this pattern^88,89^, the mediation signals from left ventricular mass and myocardial wall thickness remained robust after accounting for CVD risk factors, such as high blood pressure.

AF was primarily associated with left and right atrial traits, which emerged as putative mediators of genetic risk. The link between the left atrium and AF is consistent with previous work, which has shown that PRSs for left atrial volume were associated with AF, and with mendelian randomisation analyses supporting a causal role for left atrial dysfunction in AF development^35^. Other AF PRSs have also been previously associated with greater left atrial volume^35,69^. For the right atrium, although previous studies have suggested that right atrial and left atrial traits contribute independently to AF risk^90^, we found that the associations with right atrial traits were markedly attenuated after accounting for the other cardiac chambers, including the left atrium. Similarly, despite Mendelian randomisation evidence supported causal associations of both left and right ventricular traits with AF, our findings suggest that ventricular traits do not represent robust mediators of polygenic risk for AF^77^.

AAA was consistently associated with the descending aorta, even though some associations lost statistical significance in sensitivity analysis. However, this was likely due to reduced power from the smaller case numbers and increased model complexity with post-exposure confounding. Although cardiovascular magnetic resonance primarily captures the thoracic rather than abdominal aorta, this finding may reflect shared biology between thoracic and abdominal aneurysmal disease^91^, and is consistent with recent genetic and mendelian randomisation evidence^92^.

IS polygenic risk was reflected across all cardiac chambers; however, single-mediator analysis primarily identified left ventricular myocardial wall thickness as putative mediator. Although previous studies support a more prominent role of the left atrium in stroke risk^35,69,71^, in some cases supporting a causal effect on disease^93^, in our analyses the left ventricle appeared more robust, with left atrial traits not surviving multiple testing adjustments in single-mediator sensitivity analysis. This is consistent with prior findings linking left ventricular hypertrophy to stroke risk^67,94,95^, with evidence of genetic correlation between left ventricular wall thickness and IS^29^, and with studies supporting an independent role of the left ventricle in stroke risk^96,97^. As in CAD, this association was not fully explained by hypertension or other CVD risk factors. However, the association between left ventricular wall thickness and incident IS did not survive multiple-testing adjustment when events occurring within one year of the imaging visit were excluded. Studies with longer follow-up and higher case count are needed to validate increased left ventricular wall thickness as a mediating pathway between polygenic risk and the incident disease.

We also found that allowing CVD risk factors to lie downstream of genetic risk, while confounding the association between cardiac traits and disease, altered several associations. Considering CVD risk factors as post-exposure confounders attenuated the role of the left ventricle and the aorta in HF, strengthening that of the left atrium. While in CAD revealed a contribution of the right side of the heart. More broadly, accounting for CVD risk factors as post-exposure confounders, corroborated the evidence for indirect effects through cardiac structure across most outcomes. This pattern suggests that part of the effect of polygenic risk on cardiac traits operates through CVD risk factors. Future studies should thus investigate more explicitly the extent to which conventional risk factors mediate the relationship between polygenic risk and cardiac structure, particularly given that several of these factors, such as low-density lipoprotein cholesterol and blood pressure, are modifiable through effective and safe therapies.

From a translational perspective, our findings suggest that structural heart changes lie on a pathway from PRS to CVD, which may motivate mechanistic hypotheses and trials of cardiac remodelling-targeted prevention^98–105^. Indeed, the estimated indirect effects through cardiac structure were rather substantial for HF, IS and AF, particularly when compared to the ones of other CVDs risk factors^75,76^. However, these findings should be regarded as hypothesis generating, and studies with longer follow-up, longitudinal imaging measurements and larger number of incident cases are needed to corroborate the magnitude and robustness of these effects.

In HF, several treatments have been shown to reverse left ventricular remodelling and lead to better clinical outcomes^98–105^. In this context, PRSs may help identify individuals most likely to benefit from each class of therapy, as well as those who may warrant more frequent imaging follow-up. Moreover, the ability of PRSs to highlight anatomically localised associations, such as the descending aorta in AAA, further supports their utility in epidemiological research, beyond prediction. Indeed, integrating cardiac imaging into the established PRSs-disease framework reveals how polygenic risk manifests at the organ level, uncovers chamber- and disease-specific divergences, and identifies intermediate phenotypes that may serve as early biomarkers or therapeutic targets.

Several limitations should be acknowledged. First, the analysis was restricted to European ancestry participants from the UK Biobank imaging subset, limiting generalisability between ancestries and raising the possibility of selection bias, including collider bias^40,106^. Second, the relatively short follow-up period and low number of events across all outcomes reduced precision and may have contributed to unstable estimates, wide CIs, and loss of significance particularly when post-exposure confounders were included. Third, the lack of more detailed phenotyping prevented analysis of disease subtypes (such as HFpEF vs HFrEF) and the cross-sectional design precluded assessment of longitudinal remodelling. Finally, residual confounding and model misspecification cannot be excluded. Moreover, because our main analysis aimed to estimate conditional associations, the results should not be interpreted as marginal effects.

In conclusion, this study provides empirical evidence that PRSs for cardiovascular diseases are reflected in cardiac structure and function. Mapping the specific cardiac traits that lie on the pathway between PRSs and incident CVDs revealed distinct patterns across diseases. Crucially, these organ-level insights contribute to generate mechanistic hypotheses, highlight potential targets for early interventions, and motivate PRS-guided prevention trials for cardiac remodelling. Moving forward, our findings support future studies investigating how PRSs and CVDs risk factors jointly shape cardiac structure and function.

## Data Availability

The data generated by this study are available in the supplementary materials. Source data for the display figures has been provided. All the data fields used are reported in the supplementary materials, and requests to access UK Biobank data can be made here: www.ukbiobank.ac.uk/enable-your-research/apply-for-access.

## Acknowledgements

This research has been conducted using the UK Biobank Resource under Application Number 608471. This work was supported by core funding from the British Heart Foundation (RG/F/23/110103), NIHR Cambridge Biomedical Research Centre (NIHR203312) [*], BHF Chair Award (CH/12/2/29428), Cambridge BHF Centre of Research Excellence (RE/24/130011), and by Health Data Research UK, which is funded by the UK Medical Research Council, Engineering and Physical Sciences Research Council, Economic and Social Research Council, Department of Health and Social Care (England), Chief Scientist Office of the Scottish Government Health and Social Care Directorates, Health and Social Care Research and Development Division (Welsh Government), Public Health Agency (Northern Ireland), British Heart Foundation and the Wellcome Trust. S.R. was supported by the BHF Cambridge Centre for Research Excellence RE/24/130011. S.K. was supported by the Wellcome Trust [Grant number 220540/Z/20/A]. E.P. was supported by the British Heart Foundation (RG/F/23/110103). Y.L. was supported by The British Heart Foundation (BHF) and the German Centre for Cardiovascular Research (DZHK) (SP/F/23/150048). S.I. was supported by Cancer Research UK (EDDAPA-2024/100011). J.H.F.R. is part-supported by the EPSRC, the NIHR Cambridge Biomedical Research Centre (NIHR203312) and the British Heart Foundation Centre of Research Excellence (RE/24/130011). M.I. is supported by the Munz Chair of Cardiovascular Prediction and Prevention and the NIHR Cambridge Biomedical Research Centre (NIHR203312) [*] as well as by the UK Economic and Social Research Council (ES/T013192/1). *The views expressed are those of the authors and not necessarily those of the NIHR or the Department of Health and Social Care.

