## Supplementary materials for "Polygenic risk of cardiovascular disease manifests in cardiac structure and function"

**Supplementary Information**

**Table of Contents**

Supplementary Table Legends	2

Supplementary Figures	8

Supplementary Methods	16 Supplementary Results	18

### **Supplementary Table Legends**

**Table S1: Definitions of cardiac IDPs, covariates, and risk factors.** This table reports the UK Biobank fields corresponding to each cardiac imaging-derived phenotype (IDP) and describes the definitions used for all covariates. Cardiac IDPs, age, sex, body mass index (BMI), assessment centre, genotype array, and genetic principal components (PCs) were obtained directly from the relevant UK Biobank fields. Family history was derived by combining information from three fields, and current diabetes was defined using self-reported doctor-diagnosed diabetes. Daily cigarette consumption was recoded to include non-smokers. High cholesterol and high blood pressure were defined using information on medication use, self-reported disease, and measured blood pressure for hypertension.

**Table S2: Case definition.** This table lists how the cases for heart failure, abdominal aortic aneurysm, ischaemic stroke, coronary artery disease and atrial fibrillation were defined. To define diseases, we used International Classification of Diseases 10 (ICD-10) codes, ICD-9 codes and self-reported diseases. For coronary artery disease (CAD), we additionally used Office of Population Census and Surveys revision 4 (OPCS)-4.

**Table S3: Polygenic scores used in main and secondary analysis.** This table summarises the specific polygenic risk scores (PRSs) used in our analyses. The “PRS label” column lists the labels used throughout the tables to denote each PRS, and the “Analysis” column indicates whether the PRS was used in the main or sensitivity analyses. The “Publication and DOI” column identifies the study in which the score was originally reported, together with the corresponding digital object identifier (DOI). Finally, the “Source” column specifies whether the PRS was obtained from the PGS Catalog or UK Biobank, with the relevant links provided. PRS/PGS, polygenic risk scores.

**Table S4: Baseline characteristics.** This table reports the baseline characteristics and the values of cardiovascular disease (CVDs) risk factors, as well as the number of incident cases for the imaging sample. We reported the mean and standard deviation (SD) for continuous variables, as well as the number of cases and the percentages for binary variables.

**Table S5: Multiple-mediator analysis.** This table represents the direct, indirect, total effects and proportion mediated obtained from counterfactual multiple-mediator analysis using Cox proportional hazards model. We included as baseline covariates (basec): sex, age, BMI, daily cigarette consumption, current diabetes, current high blood pressure, current high cholesterol, family history of heart disease, assessment centre, genotype array, and the first 10 genetic PCs. We used the “cmest” function in the “CMAverse” package^1^, 1000 bootstrap resamples, and the g-formula^2^ approach. The total natural indirect effect (Rtnie), total natural direct effect (Rtnde), and total effect (Rte) were estimated on the hazard ratio (HR) scale, whereas the proportion mediated (pm) was estimated on the excess HR scale. All cardiac IDPs that were significant in the single-mediator analysis (p ≤ 0.05) were included simultaneously as putative mediators. We considered PRSs for each disease as exposure, the selected IDPs together as mediators, and the corresponding incident CVD as outcome. As all mediators were continuous IDPs, mediator models were fitted using linear regressions. We assumed no exposure-mediator interaction (Emint = F). As contrast between two specified exposure values, we set 0 as the control value and 1 as the treatment value. We used estimation = "imputation" and inference = "bootstrap". P-values were adjusted using Benjamini-Hochberg (BH) correction table-wise. Across all analysis individuals with prevalent CVDs were excluded. **Abbreviations:** PRS, polygenic risk scores; HF, heart failure; CAD, coronary artery disease; AF, atrial fibrillation; AAA, abdominal aortic aneurysm; IS, ischaemic stroke.

**Table S6: Associations between CVDs PRSs and cardiac IDPs.** This table reports the estimates of the associations between CVDs PRSs and cardiac IDPs using linear regression adjusted for sex, age, genotype array, assessment centre and first 10 genetic PCs. Both PRSs and cardiac IDPs were standardised, so the estimates represent the SD change in the imaging trait associated with a one SD increase in the PRS. The p-values were adjusted using BH correction table-wise. **Abbreviations:** CVD, cardiovascular disease; IDPs, imaging-derived phenotypes; PRS, polygenic risk scores; HF, heart failure; CAD, coronary artery disease; AF, atrial fibrillation; AAA, abdominal aortic aneurysm; IS, ischaemic stroke; LV, left ventricle; RV, right ventricle; LA, left atrium; RA, right atrium.

**Table S7: Association between cardiac IDPs and incident CVDs.** This table reports the estimates of the associations between cardiac imaging traits and the 5 incident CVDs using Cox proportional hazards models. We adjusted for sex, age, corresponding PRS, BMI, daily cigarette consumption, current high cholesterol, current high blood pressure, current diabetes, family history of CVDs and assessment centre. The estimate represents the change in the HR associated with one SD increase in cardiac IDPs. The adjusted p-value represents the p-value after the BH correction applied table-wise. **Abbreviations:** CVD, cardiovascular disease; IDPs, imaging-derived phenotypes; PRS, polygenic risk score; HF, heart failure; CAD, coronary artery disease; AF, atrial fibrillation; AAA, abdominal aortic aneurysm; IS, ischaemic stroke; LV, left ventricle; RV, right ventricle; LA, left atrium; RA, right atrium; HR, hazard ratio.

**Table S8: Indirect effects of cardiac IDPs from single-mediator analysis.** This table reports the indirect effects (expressed in HRs) obtained from counterfactual single-mediator analysis using Cox proportional hazards model. We used the “cmest” function in the “CMAverse” package^1^, 1000 bootstrap resamples and the g-formula^2^ approach. We considered PRS for each disease as exposure, each cardiac IDP as mediator and the corresponding incident CVD as outcome. We included as baseline covariates (basec): sex, age, BMI, daily cigarette consumption, current diabetes, current high blood pressure, current high cholesterol, family history of heart disease, assessment centre, genotype array, and the first 10 genetic PCs. Since all mediators were continuous IDPs, mediator models were fitted using linear regression. We assumed no exposure-mediator interaction (Emint = F). As contrast between two specified exposure values, we set 0 as the control value and 1 as the treatment value. We used estimation = "imputation" and inference = "bootstrap". P-values were adjusted using BH correction table-wise. **Abbreviations:** IDPs, imaging-derived phenotypes; PRS, polygenic risk score; HF, heart failure; CAD, coronary artery disease; AF, atrial fibrillation; AAA, abdominal aortic aneurysm; IS, ischaemic stroke; LV, left ventricle; RV, right ventricle; LA, left atrium; RA, right atrium.

**Table S9: Indirect effects from single-mediator analyses adjusted for all other cardiac chambers, except the chamber corresponding to the trait of interest.** This table reports the indirect effects (expressed in HR) derived from the single-mediator analysis, adjusting for all cardiac chambers except the chamber corresponding to trait of interest (e.g., adjusting for PCs representing the left atrium, right atrium, right ventricle and aorta when analysing left ventricular traits). Therefore, in Cox proportional hazards models, we considered the following variables as baseline covariates (basec): sex, age, BMI, daily cigarette consumption, current diabetes, current high blood pressure, current high cholesterol, family history of heart disease, assessment centre, genotype array, the first 10 genetic PCs and PCs from all the other cardiac chambers except the one corresponding to the trait of interest. (**Supplementary methods**). P-values were adjusted using BH correction table-wise. The remaining specifications for the single-mediator analysis were the same as those reported in **Table S8**. **Abbreviations:** IDPs, imaging-derived phenotypes; PRS, polygenic risk score; HF, heart failure; CAD, coronary artery disease; AF, atrial fibrillation; AAA, abdominal aortic aneurysm; IS, ischaemic stroke; LV, left ventricle; RV, right ventricle; LA, left atrium; RA, right atrium.

**Table S10: Single-mediator analysis for left ventricular traits in heart failure, accounting for left ventricular ejection fraction.** To understand if the mediation signals for some left ventricular traits were driven by the left ventricular ejection fraction, we accounted for it in single-mediator analysis both as post-exposure confounder and as baseline covariate. In the first case, we assumed that genetic risk initially influenced left ventricular ejection fraction, which in turn influenced the other left ventricular traits. We therefore modelled left ventricular ejection fraction as post-exposure confounder using a linear regression (postc). In the second case, we considered left ventricular ejection fraction as baseline covariate (basec), adjusting for it in the Cox proportional hazards model beyond sex, age, BMI, current high cholesterol, current high blood pressure, current diabetes, daily cigarette consumption, family history of CVDs, genotype array, assessment centre and first 10 genetic PCs. P-values were adjusted using Benjamini-Hochberg (BH) procedure to control for multiple testing for each table separately. The remaining specifications for the single-mediator analysis were the same as those reported in **Table S8**. We also repeated the same analysis using the alternative set of PRSs (**Table S3**). **Abbreviations:** IDPs, imaging-derived phenotypes; PRS, polygenic risk score; HF, heart failure; LV, left ventricle; LVEF, left ventricular ejection fraction.

**Table S11: Associations between PRSs and cardiac IDPs adjusting for established risk factors.** This table reports the results of the association between CVDs PRSs and cardiac IDPs, adjusting for BMI, daily cigarette consumption, current high cholesterol, current high blood pressure, current diabetes and family history of CVDs, beyond sex, age, genotype array, assessment centre and first 10 genetic PCs. Both PRSs and imaging traits were standardised, so the estimates represent the SD change in the imaging trait associated with a one SD increase in the PRS. The p-values were adjusted using BH procedure to control for multiple testing table-wise. **Abbreviations:** IDPs, imaging-derived phenotypes; PRS, polygenic risk score; HF, heart failure; CAD, coronary artery disease; AF, atrial fibrillation; AAA, abdominal aortic aneurysm; IS, ischaemic stroke; LV, left ventricle; RV, right ventricle; LA, left atrium; RA, right atrium.

**Table S12: Association of atrial fibrillation PRSs and the Townsend deprivation index (TDI) with left ventricular wall thickness.** This table reports the results of the association of left ventricular wall thickness with atrial fibrillation (AF) PRSs and the Townsend deprivation index. This analysis was aimed to understand how genetic (AF PRS) and environmental factors (Townsend deprivation index - TDI) were associated with left ventricular wall thickness. We used a linear regression model, considering left ventricular wall thickness as the outcome and adjusting for the TDI (Data-Field 22189), AF PRS, age, sex, assessment centre, genotype array, and the first 10 genetic PCs. Regression coefficients, standard errors, and p-values are reported. **Abbreviations:** AF, atrial fibrillation.

**Table S13: Indirect effects from single-mediator analysis considering BMI, high cholesterol, high blood pressure and diabetes as post-exposure confounders.** This table represents the indirect effects (expressed in HRs) obtained from single-mediator analysis modelling BMI, current high cholesterol, current high blood pressure and current diabetes as post-exposure confounders, namely as confounders of cardiac IDPs and CVDs affected by the PRSs (considering them as postc). BMI was modelled using a linear regression while binary variables, namely current high cholesterol, current high blood pressure, and current diabetes were modelled using logistic regression. Therefore, in Cox proportional hazards model, we considered the following variables as baseline covariates (basec): sex, age, daily cigarette consumption, family history of heart disease, assessment centre, genotype array, the first 10 genetic PCs. P-values were adjusted using BH procedure to control for multiple testing table-wise. The remaining specifications for the single-mediator analysis were identical to those reported in **Table S8**, except that 200 bootstrap iterations were used for this sensitivity analysis. **Abbreviations:** BMI, body mass index; IDPs, imaging-derived phenotypes; PRS, polygenic risk score; HF, heart failure; CAD, coronary artery disease; AF, atrial fibrillation; AAA, abdominal aortic aneurysm; IS, ischaemic stroke; LV, left ventricle; RV, right ventricle; LA, left atrium; RA, right atrium.

**Table S14: Sensitivity analyses for multiple-mediator analyses.** This table represents the results from multiple-mediator analysis implementing several sensitivity analyses: (1) Considering BMI, current high blood pressure, current high cholesterol and current diabetes as post-exposure confounders, (2) using the alternative set of PRSs for each disease (**Table S3**), (3) using exposure contrast from the right part of the PRSs distribution, (4) using exposure contrast from the left part of the PRSs distribution, (5) assuming exposure-mediator interaction, (6) using all IDPs as mediators without preselecting them, (7) using as mediators only the IDPs which were highly significant in single-mediator analysis (q≤0.05) (**Table S8**). P-values were adjusted using BH procedure to control for multiple testing for each table separately. The remaining specifications for the multiple-mediator analyses were identical to those reported in **Table S5**. **Abbreviations:** IDPs, imaging-derived phenotypes; PRS, polygenic risk score; HF, heart failure; CAD, coronary artery disease; AF, atrial fibrillation; AAA, abdominal aortic aneurysm; IS, ischaemic stroke; LV, left ventricle; RV, right ventricle; LA, left atrium; RA, right atrium.

**Table S15: Associations between PRSs and cardiac IDPs excluding also incident cases.** This table reports the estimates of the association between CVDs PRSs and cardiac IDPs excluding incident cases, to minimise the risk of reverse causation. We used linear regression adjusted for sex, age, genotype array, assessment centre and first 10 genetic PCs. Both PRSs and imaging traits were standardised, so the estimates represent the SD change in the imaging trait associated with a one SD increase in the PRS. The p-values were adjusted using BH procedure to control for multiple testing table-wise. **Abbreviations:** IDPs, imaging-derived phenotypes; PRS, polygenic risk score; HF, heart failure; CAD, coronary artery disease; AF, atrial fibrillation; AAA, abdominal aortic aneurysm; IS, ischaemic stroke; LV, left ventricle; RV, right ventricle; LA, left atrium; RA, right atrium.

**Table S16: Associations between PRSs and cardiac IDPs using alternative PRSs.** This table presents the results of the association between CVDs PRSs and cardiac IDPs using the alternative PRSs for each disease (**Table S3**). We used linear regression adjusted for sex, age, genotype array, assessment centre and first 10 genetic PCs. Both PRSs and imaging traits were standardised, so the estimates represent the SD change in the imaging trait associated with a one SD increase in the PRS. The p-values were adjusted using BH procedure to control for multiple testing table-wise. **Abbreviations:** IDPs, imaging-derived phenotypes; PRS, polygenic risk score; HF, heart failure; CAD, coronary artery disease; AF, atrial fibrillation; AAA, abdominal aortic aneurysm; IS, ischaemic stroke; LV, left ventricle; RV, right ventricle; LA, left atrium; RA, right atrium.

**Table S17: Association between cardiac IDPs and incident diseases excluding cases occurring within one year of the imaging visit.** This table reports the results of the association between cardiac IDPs and the 5 incident CVDs, excluding individuals who developed the disease within one year of the imaging visit, to minimise the risk of reverse causation. We used Cox proportional hazards model adjusting for sex, age, corresponding PRS, BMI, current high cholesterol, current high blood pressure, current diabetes, daily cigarette consumption, family history of CVDs and assessment centre. The “HR” column represents the change in the HR associated with one SD increase in cardiac IDPs. The p-values were adjusted using BH procedure to control for multiple testing table-wise. **Abbreviations:** IDPs, imaging-derived phenotypes; PRS, polygenic risk score; HF, heart failure; CAD, coronary artery disease; AF, atrial fibrillation; AAA, abdominal aortic aneurysm; IS, ischaemic stroke; LV, left ventricle; RV, right ventricle; LA, left atrium; RA, right atrium; HR, hazard ratio.

**Table S18: Indirect effects from single-mediator analysis** **using the alternative PRS for each disease.** This table reports the results of a sensitivity analysis for single-mediator analysis using the alternative set of PRSs as exposures (**Table S3**). The specifications used were identical to those reported in the main single-mediator analysis in **Table S8**. The indirect effects are expressed in HRs and P-values were adjusted using BH procedure to control for multiple testing table-wise. 200 bootstrap iterations were used for this sensitivity analysis. **Abbreviations:** IDPs, imaging-derived phenotypes; PRS, polygenic risk score; HF, heart failure; CAD, coronary artery disease; AF, atrial fibrillation; AAA, abdominal aortic aneurysm; IS, ischaemic stroke; LV, left ventricle; RV, right ventricle; LA, left atrium; RA, right atrium.

**Table S19: Indirect effects from single-mediator analysis** **using an exposure contrast from the right part of PRSs distribution.** This table presents the indirect effects (expressed in HRs) obtained from single-mediator analysis using an alternative exposure contrast, to assess the robustness of findings. The contrast between exposure values was set to 3, as the control value, and to 4, as the treatment value, namely a contrast from the higher part of the PRS distribution. P-values were adjusted using BH procedure to control for multiple testing table-wise. The remaining specifications for the single-mediator analysis were identical to those reported in **Table S8**, except that 200 bootstrap iterations were used for this sensitivity analysis. **Abbreviations:** IDPs, imaging-derived phenotypes; PRS, polygenic risk score; HF, heart failure; CAD, coronary artery disease; AF, atrial fibrillation; AAA, abdominal aortic aneurysm; IS, ischaemic stroke; LV, left ventricle; RV, right ventricle; LA, left atrium; RA, right atrium.

**Table S20: Indirect effects from single-mediator analysis** **using an exposure contrast from the left part of PRSs distribution.** This table presents the indirect effects in HR obtained from single-mediator analysis using an alternative exposure contrast, to assess the robustness of results. The contrast between exposure values was set to -4, as the control value, and to -3, as the treatment value, namely a contrast from the lower part of the PRS distribution. P-values were adjusted using BH procedure to control for multiple testing table-wise. The remaining specifications for the single-mediator analysis were identical to those reported in **Table S8**, except that 200 bootstrap iterations were used for this sensitivity analysis. **Abbreviations:** IDPs, imaging-derived phenotypes; PRS, polygenic risk score; HF, heart failure; CAD, coronary artery disease; AF, atrial fibrillation; AAA, abdominal aortic aneurysm; IS, ischaemic stroke; LV, left ventricle; RV, right ventricle; LA, left atrium; RA, right atrium.

**Table S21: Indirect effects from single-mediator analysis assuming exposure-mediator interaction.** The table represents the indirect effects (expressed in HRs) obtained from single-mediator analysis assuming exposure-mediator interaction (Emint = T), to assess the robustness of results. P-values were adjusted using BH procedure to control for multiple testing table-wise. The remaining specifications for the single-mediator analysis were identical to those reported in **Table S8**, except that 200 bootstrap iterations were used for this sensitivity analysis. **Abbreviations:** IDPs, imaging-derived phenotypes; PRS, polygenic risk score; HF, heart failure; CAD, coronary artery disease; AF, atrial fibrillation; AAA, abdominal aortic aneurysm; IS, ischaemic stroke; LV, left ventricle; RV, right ventricle; LA, left atrium; RA, right atrium.

**Table S22: PCs extracted separately from each cardiac chamber.** This table reports results from principal component analysis (PCA), performed separately for left ventricular, left atrial, right atrial, right ventricular, and aortic traits using the “prcomp” function in R. For each PC within each chamber, the SD, proportion of variance explained, cumulative proportion of variance explained, and loadings are provided. **Abbreviations:** PC, principal component; imaging-derived phenotypes; LV, left ventricle; RV, right ventricle; LA, left atrium; RA, right atrium; Ao, aorta.

**Table S23: Analyses using AF PRS without PITX2 gene.** Given that the direction of the associations between AF PRS and left ventricular myocardial mass as well as left ventricular wall thickness were unexpected, we hypothesised that these associations were driven by the PITX2 gene, which is already known to play an important role in AF (**Supplementary methods**). Therefore, we repeated our analyses using a new AF PRS, not containing the Single Nucleotide Polymorphisms (SNPs) associated with PITX2 gene (**Supplementary methods**). First, we assessed the association between AF PRSs and IDPs considering the new AF PRS, and using the same specifications reported in **Table S6**. Second, we implemented single-mediator analysis using the new AF PRS and the same specifications reported in **Table S8**, except that 200 bootstrap iterations were used for this sensitivity analysis. P-values were adjusted using BH procedure to control for multiple testing for each table separately. **Abbreviations:** IDPs, imaging-derived phenotypes; PRS, polygenic risk score; LV, left ventricle; RV, right ventricle; LA, left atrium; RA, right atrium.

**Table S24: Assessment of Cox proportional hazards models which violated the proportional hazards assumption using time interaction analysis.** This table presents the results of Cox proportional hazards model including a time interaction term for the cardiac imaging traits. We ran this sensitivity analysis for the models which violated the proportional hazard**s** assumption. We reported the HR both for the cardiac IDPs and the time interaction term with corresponding standard error and p-value. **Abbreviations:** PH, proportional hazards; HF, heart failure; CAD, coronary artery disease; LV, left ventricle; LA, left atrium; HR, hazard ratio.

**Table S25: Single-mediator analysis adjusting for all cardiac chambers except the one corresponding to the trait of interest, using the alternative set of PRSs.** Similarly to **Table S9**, this table reports the indirect effects (expressed in HRs) derived from the single-mediator analysis, adjusted for all cardiac chambers except the one corresponding to the trait of interest, thereby providing conditional estimates. The difference with **Table S9** is that in this case we used the alternative set of PRSs **(Table S3).** In Cox proportional hazards model, we considered the following variables as baseline covariates (basec): sex, age, BMI, daily cigarette consumption, current diabetes, current high blood pressure, current high cholesterol, family history of heart disease, assessment centre, genotype array, the first 10 genetic PCs and PCs from all the other cardiac chambers except the one corresponding to the trait of interest. P-values were adjusted using BH procedure to control for multiple testing table-wise. The remaining specifications for the single-mediator analysis were identical to those reported in **Table S8**, except that 200 bootstrap iterations were used for this sensitivity analysis. **Abbreviations:** IDPs, imaging-derived phenotypes; PRS, polygenic risk score; HF, heart failure; CAD, coronary artery disease; AF, atrial fibrillation; AAA, abdominal aortic aneurysm; IS, ischaemic stroke; LV, left ventricle; RV, right ventricle; LA, left atrium; RA, right atrium.

**Table S26: Single-mediator analysis** **of heart failure, coronary artery disease and ischaemic stroke accounting for the left ventricle.** Given the already established relevance of the left ventricle in heart failure, coronary artery disease and ischaemic stroke (**Supplementary methods**), we decided to account for it both as post-exposure confounder and as baseline covariate. In the first case, we assumed that genetic risk influenced initially left ventricular traits which in turn influenced the remaining cardiac structures, therefore we modelled PCs extracted from left ventricular traits as post-exposure confounders using linear regressions (considered as postc). In the second case, instead, we modelled PCs extracted from left ventricular traits as baseline covariates (considered as basec). Adjusting for them beyond sex, age, BMI, current high cholesterol, current high blood pressure, current diabetes, daily cigarette consumption, family history of CVDs, genotype array, assessment centre and first 10 genetic PCs. Both analyses were aimed to understand if the mediation signals from other chambers, beyond the left ventricle, remained even after accounting for it. P-values were adjusted using BH procedure to control for multiple testing considering each table separately. The remaining specifications for the single-mediator analysis were the same as those reported in **Table S8**, except that 200 bootstrap iterations were used for this sensitivity analysis. **Abbreviations:** IDPs, imaging-derived phenotypes; PRS, polygenic risk score; HF, heart failure; CAD, coronary artery disease; IS, ischaemic stroke; LV, left ventricle; RV, right ventricle; LA, left atrium; RA, right atrium.

**Table S27: Single-mediator analysis** **using alternative PRSs for heart failure, coronary artery disease and ischaemic stroke and accounting for the left ventricle.** As in **Table S26**, this table presents the estimates from single-mediator analysis accounting for the left ventricle, both as post-exposure confounder and as baseline covariates. The difference with **Table S26** is that this time we used the alternative set of PRSs (**Table S3**). The remaining specifications were identical to those reported in **Table S26**. **Abbreviations:** IDPs, imaging-derived phenotypes; PRS, polygenic risk score; HF, heart failure; CAD, coronary artery disease; IS, ischaemic stroke; LV, left ventricle; RV, right ventricle; LA, left atrium; RA, right atrium.

**Table S28: Single-mediator analysis** **for atrial fibrillation and ischaemic stroke accounting for the left atrium.** Given the already established relevance of the left atrium in atrial fibrillation and ischaemic stroke (**Supplementary information about methods**), we decided to account for it both as post-exposure confounder and as baseline covariate. In the first case, we assumed that genetic risk influenced initially left atrial traits which in turn influenced the remaining cardiac structures, therefore we modelled PCs extracted from left atrial traits as post-exposure confounders using linear regressions (considered as postc). In the second case, instead, we modelled the PCs extracted from left atrial traits as baseline covariates (considered as basec), adjusting for them beyond sex, age, BMI, current high cholesterol, current high blood pressure, current diabetes, daily cigarette consumption, family history of CVDs, genotype array, assessment centre and first 10 genetic PCs. Both analyses were aimed to understand if the mediation signals from other chambers, beyond the left atrium, remained even after accounting for it. The remaining specifications for the single-mediator analysis were the identical to those reported in **Table S8**, except that 200 bootstrap iterations were used for this sensitivity analysis. We did the same also using the alternative set of PRSs (**Table S3**). P-values were adjusted using BH procedure to control for multiple testing considering each table separately. **Abbreviations:** IDPs, imaging-derived phenotypes; PRS, polygenic risk score; AF, atrial fibrillation; IS, ischaemic stroke; LV, left ventricle; RV, right ventricle; LA, left atrium; RA, right atrium.

### **Supplementary Figures**

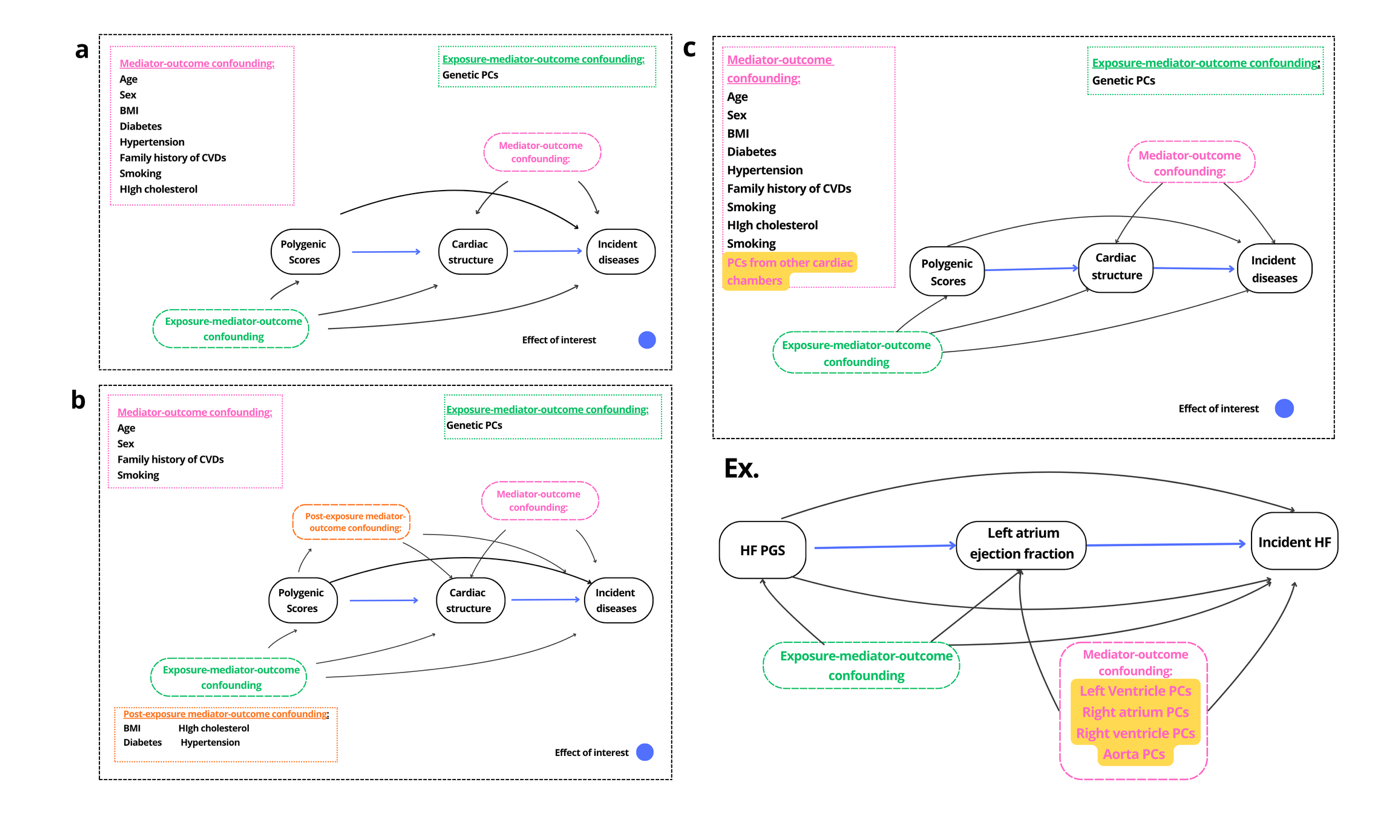

**Supplementary Figure S1. Directed acyclic graph (DAG) for mediation analysis**

**Panel (a)** presents the DAG for our main analysis. Mediator-outcome confounding is highlighted in pink, exposure-mediator-outcome confounding in green, and the effect of interest in blue, namely indirect effects.

**Panel (b)** presents the DAG for sensitivity analysis. It differs from the main analysis by the presence of post-exposure mediators-outcome confounding, highlighted in orange, representing variables that affect both cardiac structure and CVD outcomes, and that are themselves influenced by the CVDs polygenic scores.

**Panel (c)** presents the DAG used to account for cardiac chambers, other than the one corresponding to the trait of interest, considering them as baseline covariates (in pink and highlighted in yellow). This analysis therefore estimates the indirect effect of each cardiac trait conditional on measures from the other cardiac chambers, allowing us to assess whether the mediation signals reflect correlations with other cardiac chambers.

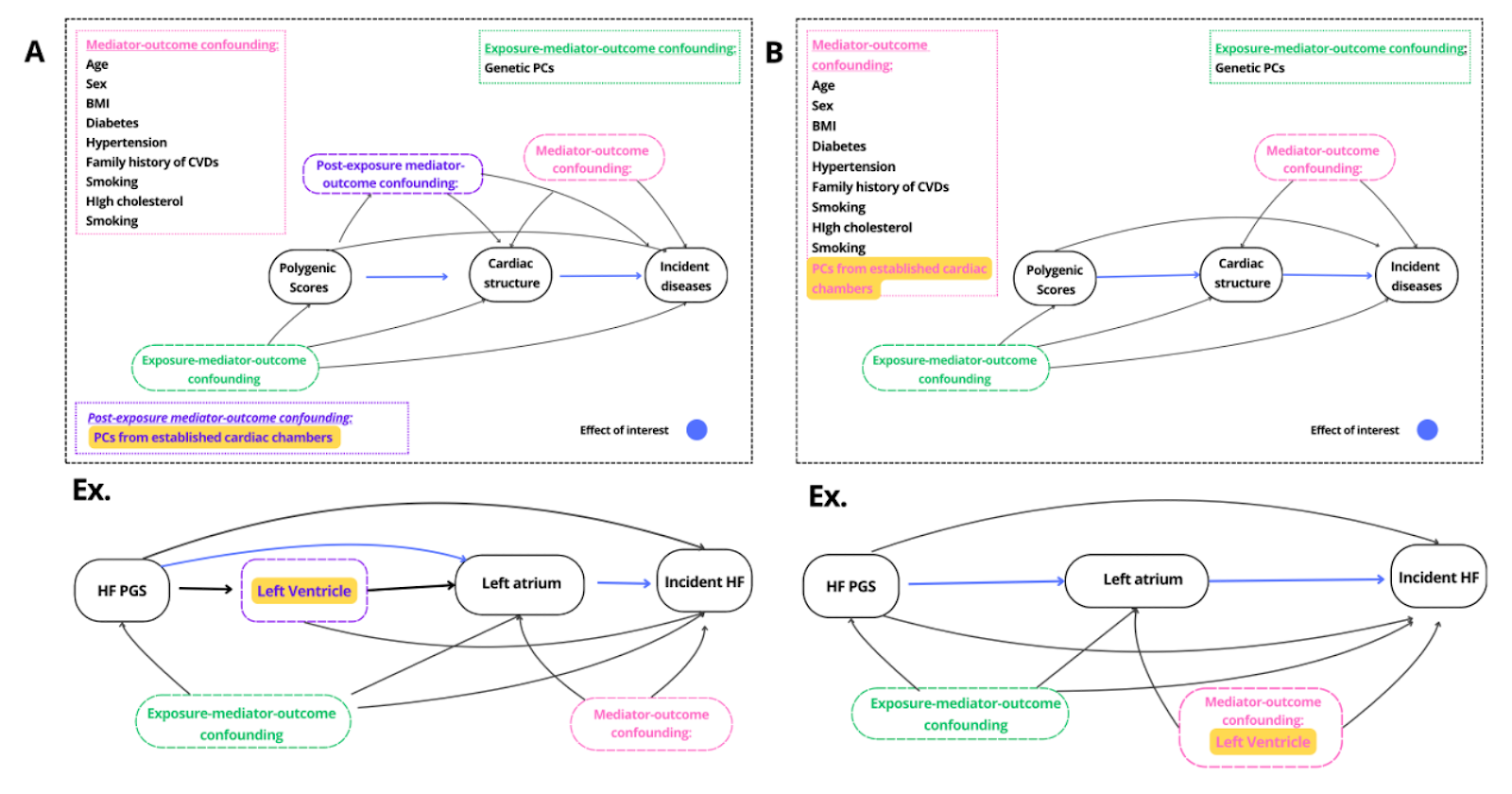

**Supplementary Figure S2: Directed acyclic graphs (DAG) used to account for established chambers in single mediation analysis**

Both panels show the assumptions used to assess whether established cardiac chambers account for the observed mediation signals. These assumptions were used to implement sensitivity analysis.

**Panel (a)** shows the DAG for the analysis which considered established cardiac chambers, such as the left ventricle in heart failure, as post-exposure mediator-outcome confounders (highlighted in purple).

**Panel (b)** shows the DAG for the analysis which considered established cardiac chambers as baseline covariates (highlighted in pink), instead of post-exposure confounders. This analysis therefore estimates the indirect effect of cardiac traits conditional on established cardiac chamber measures.

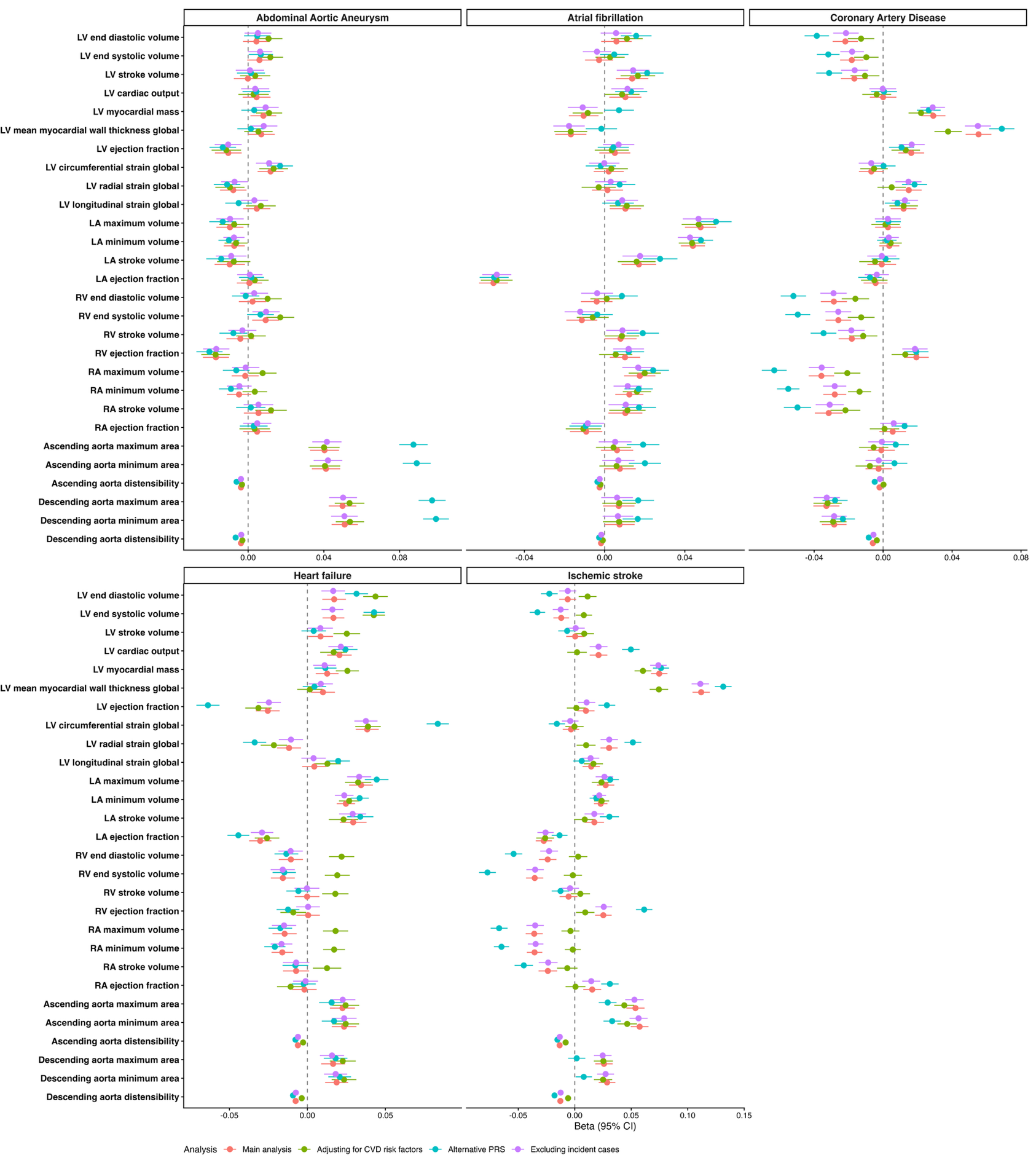

### **Supplementary Figure S3: Sensitivity analyses for the association between PRS and cardiac IDPs**

This forest plot compares the associations between PRSs and cardiac IDPs across different analyses, including the main analysis, the model further adjusting for CVD risk factors (BMI, high cholesterol, high blood pressure and diabetes), the model using the alternative PRS for the same disease, the analysis excluding also incident cases. For the main model we used robust linear regression adjusted for sex, age, first 10 genetic PCs, assessment centre and genotyping array. The beta estimates represent the SD change in imaging traits per SD increase in PRS. PGS Catalog IDs: Abdominal Aortic Aneurysm - PGS001784, Atrial fibrillation- PGS005072, Heart Failure - PGS001790. UKBB Fields IDs: ischaemic stroke (Field 26248), coronary artery disease (Field 26227).

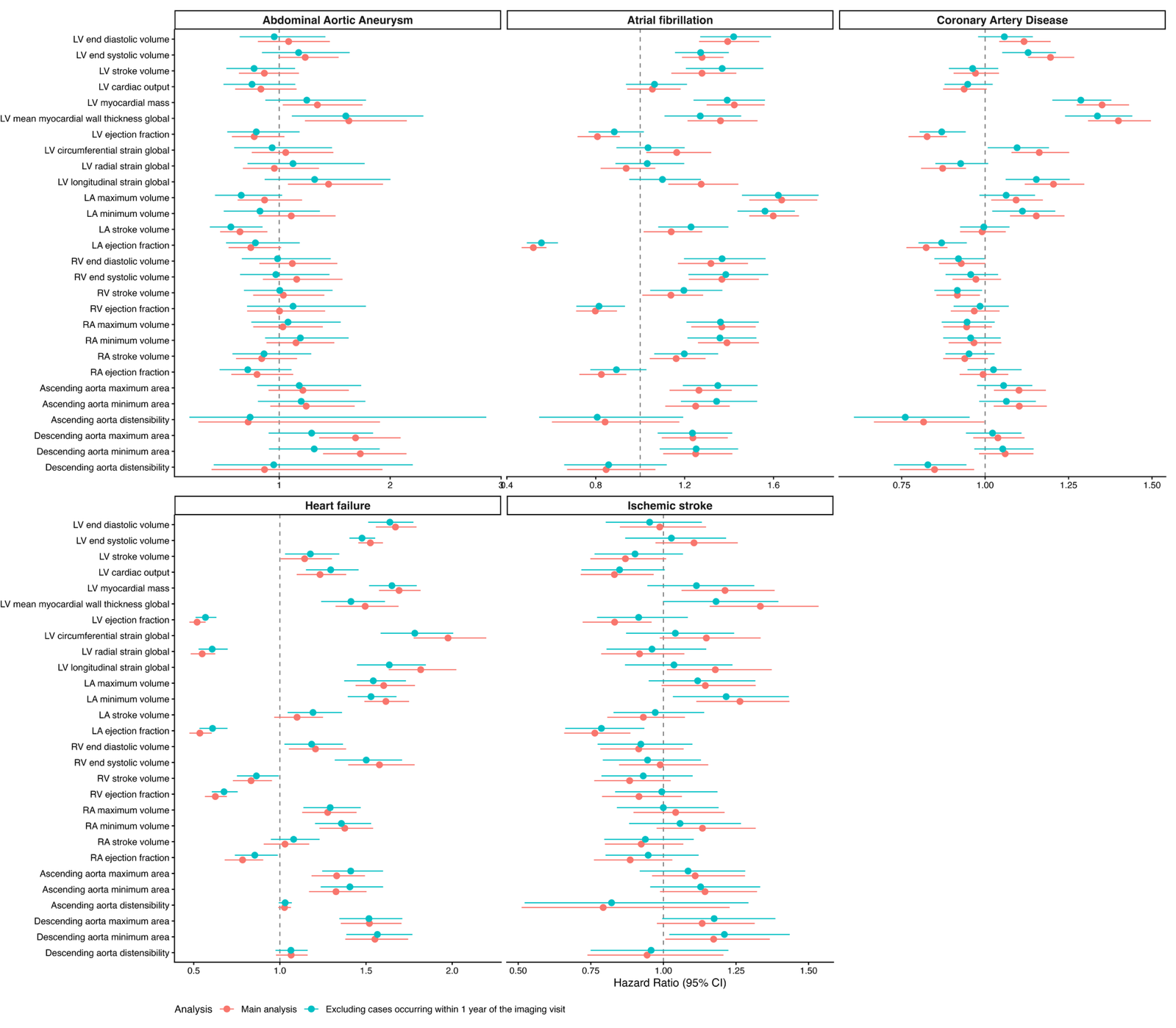

### **Supplementary Figure S4: Sensitivity analyses for the association between cardiac IDPs and CVDs**

This forest plot represents the HRs, and corresponding 95% confidence intervals, of the main analysis compared to the model excluding cases happening within one year of the imaging visit. We used in both cases Cox proportional hazards models adjusting for sex, age, corresponding PRS, daily cigarette consumption, current diabetes, current high blood pressure, current high cholesterol, BMI, family history of diseases and assessment centre.

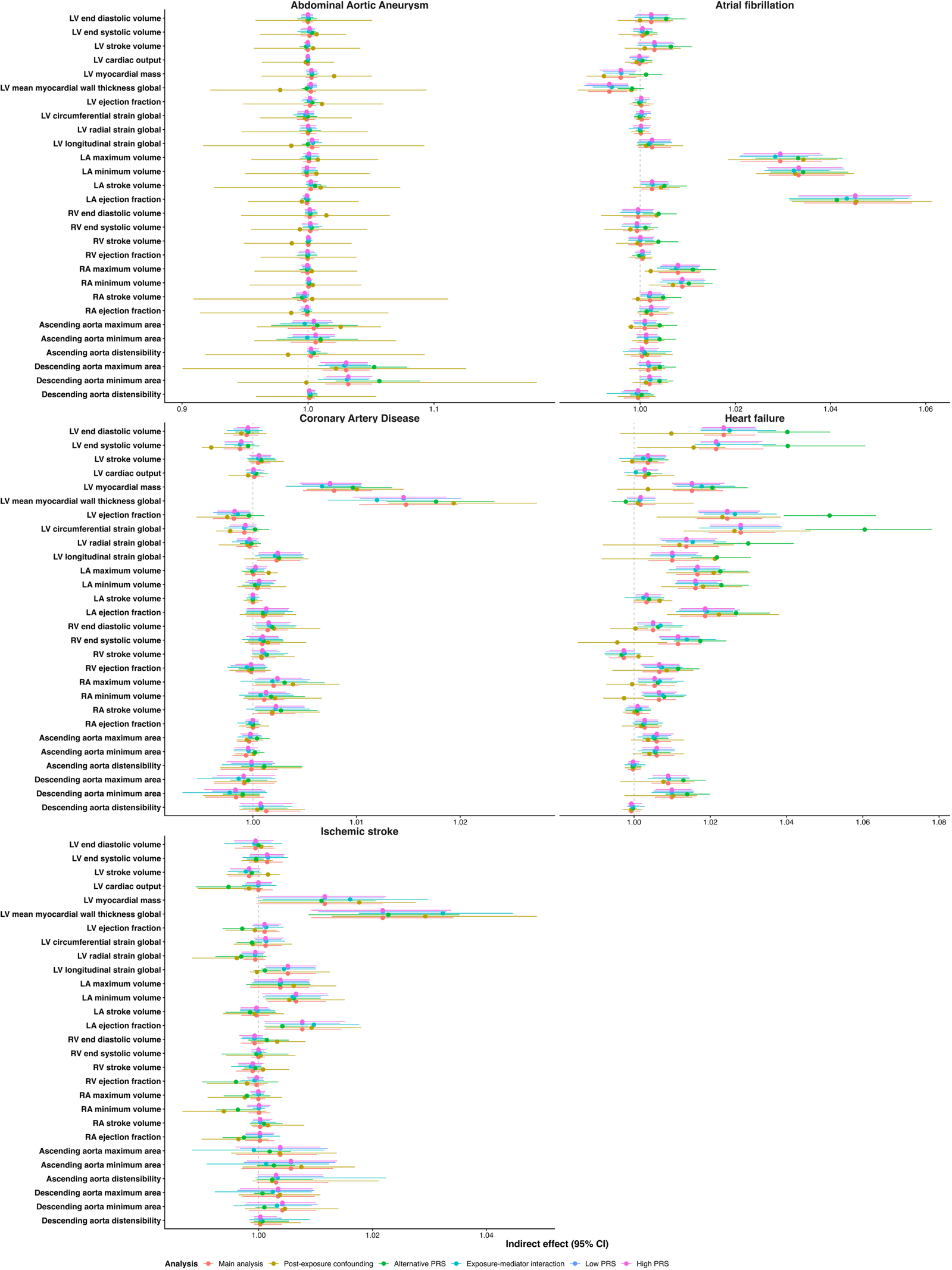

### **Supplementary Figure S5: Sensitivity analyses for single-mediator analyses**

Forest plot comparing the single-mediator indirect effects across models: (1) the main analysis, (2) model accounting for CVD risk factors as post-exposure confounders, (3) analysis using alternative disease-specific PRSs, (4) model assuming exposure-mediator interaction, (5) analysis comparing values from the upper and (6) lower part of the PRS distribution. Indirect effects are in HR from counterfactual single-mediator analysis (using Cox proportional hazards models and g-formula in “CMAverse” package). For each disease, the exposure was the PRS, the potential mediator was each cardiac trait, and the outcome was the corresponding incident disease. All models, except for the one accounting for post-exposure confounding, were adjusted for sex, age, the first 10 genetic PCs, assessment centre, genotyping array, daily cigarette consumption, current diabetes, current high blood pressure, current high cholesterol, BMI and family history of disease. The model accounting for post-exposure confounding was adjusted for sex, age, the first 10 genetic PCs, assessment centre, genotyping array, daily cigarette consumption and family history of disease while current diabetes, current high blood pressure, current high cholesterol and BMI were modelled as post-exposure confounders. CIs were derived using 1000 bootstrap iterations.

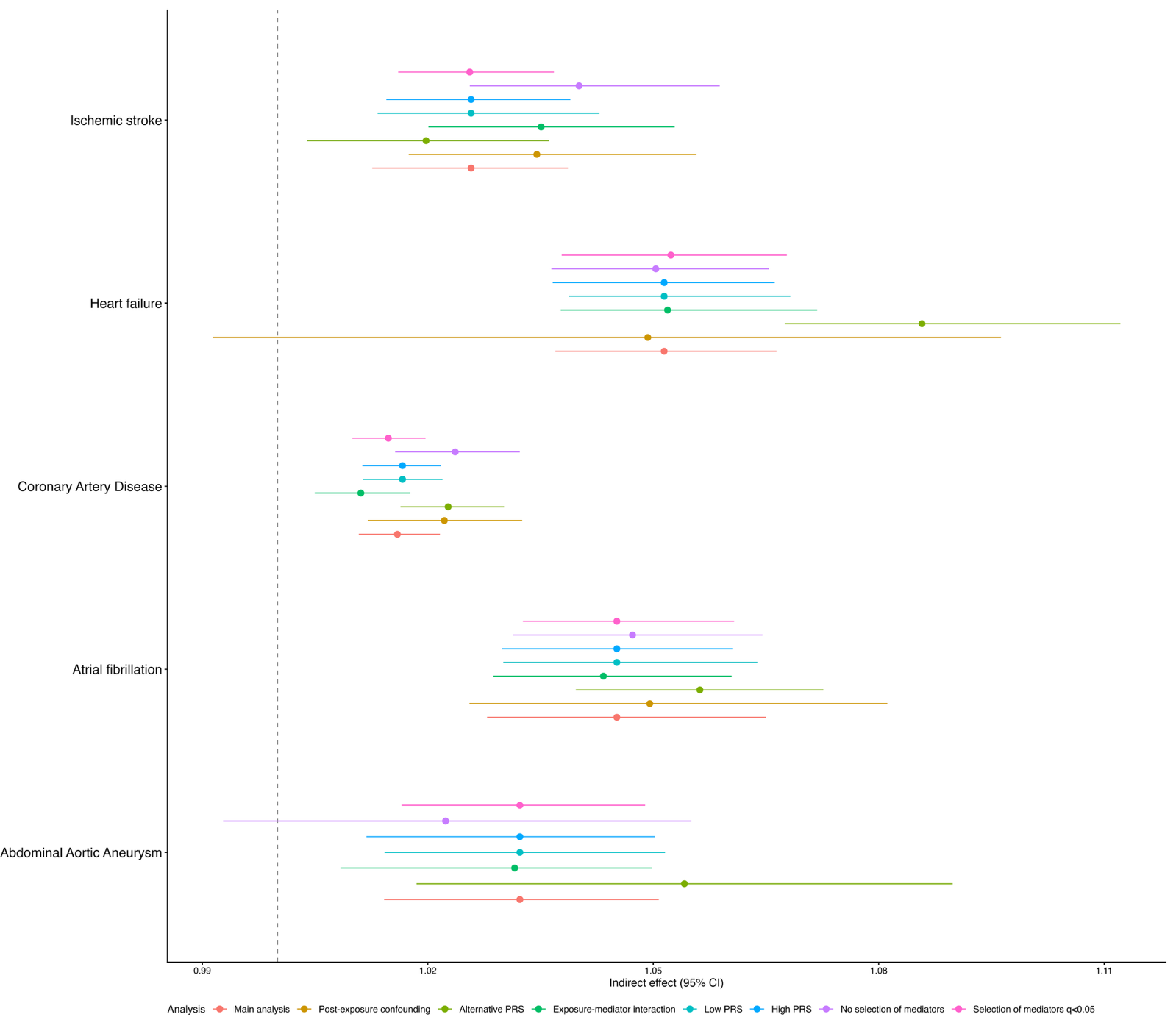

### **Supplementary Figure S6: Sensitivity analyses for multiple-mediator analyses**

Forest plot comparing the multiple-mediator indirect effects across models: (1) the main analysis, (2) model accounting for CVD risk factors as post-exposure confounders, (3) analysis using alternative disease-specific PRSs, (4) model assuming exposure-mediator interaction, (5) analysis comparing values from the upper and (6) lower part of the PRS distribution, (7) model including all cardiac IDPs without preselecting them, (8) analysis including only the cardiac IDPs with FDR ≤ 0.05 in single-mediator analysis. Indirect effects are in HR from counterfactual multiple-mediator analysis (using Cox proportional hazards models and g-formula in “CMAverse” package). For each disease, the exposure was a PRS and the outcome was the corresponding incident disease. For all models except for (7) and (8) the potential mediators were the cardiac traits significant in single-mediator analysis (p≤ 0.05). All models, except for the one accounting for post-exposure confounding, were adjusted for sex, age, the first 10 genetic PCs, assessment centre, genotyping array, daily cigarette consumption, current diabetes, current high blood pressure, current high cholesterol, BMI and family history of disease. The model accounting for post-exposure confounding was adjusted for sex, age, the first 10 genetic PCs, assessment centre, genotyping array, daily cigarette consumption and family history of disease while current diabetes, current high blood pressure, current high cholesterol and BMI were modelled as post-exposure confounders. CIs were derived using 1000 bootstrap iterations.

**Supplementary Methods**

**Counterfactual mediation analysis**

To assess the extent that cardiac structure accounted for the association between polygenic risk and incident diseases we conducted mediation analysis. We used the counterfactual framework to decompose the association between PRSs and incident CVD into components operating through cardiac structure (indirect effects) and components operating through other pathways. Let A denote the polygenic risk score, M the cardiac imaging trait, and Y the incident disease outcome. For continuous exposures, mediation effects are defined by contrasting two specified exposure values, denoted a and a*, where a* is the reference value and a is the comparison value. The total causal effect compares the outcomes that would be observed if the same individual had exposure level A = a with the outcome that would be observed if that individual instead had A = a*^3^.

The total natural direct effect represents the part of the association between PRSs and incident CVD that does not operate through cardiac structure. It compares the outcome that would be observed if an individual had exposure level A=a with the outcome that would be observed if the same individual instead had exposure level A=a*; while holding the mediator fixed in both scenarios at the value it would naturally take under A=a*^4^. On the other hand, the total natural indirect effect represents the part of the association that operates through cardiac structure. It compares the outcome that would be observed if the mediator took the value it would naturally have under A=a with the outcome that would be observed if the mediator instead took the value it would naturally have under A=a*; while the exposure is held fixed at A=a^4^.

Mediation analysis relies on four main assumptions: (1) no unmeasured confounding on the exposure-outcome relationship, (2) no unmeasured confounding on the mediator-outcome relationship, (3) no unmeasured confounding on the exposure-mediator relationship, (4) no mediator-outcome confounder influenced by the exposure^5^. We sought to make assumptions 1-3 more plausible by conditioning on the measured covariates in **Supplementary Figure S1a**; however, residual confounding from unmeasured factors cannot be ruled out. The fourth assumption was examined in sensitivity analyses (**Supplementary Figure S1b)**. In our setting, post-exposure confounders were variables that affected both cardiac structure and incident CVD, but that might also be influenced by CVD PRSs (**Supplementary Figure S1b)**. In this case, considering them merely as baseline covariates may remove part of the genetic effect we aimed to decompose.

**Single-mediator analysis adjusting for the other cardiac chambers**

We aimed to understand if the identified mediators were driven by other cardiac chambers, or if they remained robust after accounting for them. In the main analysis, we adjusted for the other cardiac chambers except the one corresponding to trait of interest, considering them as baseline covariates (e.g., adjusting for PCs extracted from the left ventricle, right atrium, right ventricle and aorta when analysing left atrial traits) (**Supplementary Figure S1c**). To do this, we derived PCs separately for each chamber, more details later in the **Supplementary methods**.

However, given the complex interrelationships among cardiac traits, some structures may act as colliders or mediators. Adjusting for such variables could therefore introduce collider bias or overadjustment bias. For this reason, we also assessed the robustness of our findings using a more restricted adjustment strategy, that included only established chambers, considering them both as post-exposure confounders and as baseline covariates. Prior literature supports a role for the left ventricle in heart failure^6–9^and in coronary artery disease^10,11^, and a more established role for the left atrium in atrial fibrillation^12,13^ and in ischaemic stroke^14,15^.

We sought to determine whether the mediation signals observed in our analyses were primarily driven by these cardiac chambers, already known to be involved, or whether mediation through other chambers remained after accounting for them. We refer to the established disease-relevant chambers as primary chambers, and to the remaining chambers as secondary chambers. We decide to account for primary chambers as post-exposure confounders in the first sensitivity analysis and to consider them as baseline covariates in second sensitivity analysis (e.g., when analysing left atrial traits in heart failure we accounted for PCs extracted from the left ventricle both as post-exposure confounders and as baseline covariates) (**Supplementary Figure S2**). In the first case, when primary chambers were included as post-exposure confounders, they were explicitly allowed to be influenced by genetic risk and to affect both secondary chambers and disease risk (**Supplementary Figure S2**). This specification therefore assessed whether secondary chambers retained explanatory value beyond the primary ones. By contrast, in the second analysis, we evaluated whether secondary chambers continued to explain the association between genetic risk and incident CVD among individuals with comparable values of the primary chambers, thereby estimating conditional effects.

**Chamber specific PCs**

To implement the previous analysis, we need to derive chamber-specific PCs, which can be considered as baseline covariates in main analysis, and both as baseline covariates and as post-exposure confounders in sensitivity analysis. PCA was performed separately for left ventricular, left atrial, right atrial, right ventricular, and aortic traits, using the “prcomp” function in R. For each PC within each chamber, the SD, proportion of variance explained, cumulative proportion of variance explained, and trait loadings are provided in **Table S22**. In the left atrium, right atrium, and right ventricle, the first PC explained approximately 60% of the variance, while the first two PCs, taken together, explained more than 97%. Because the conventional criterion of retaining components explaining around 80% of the variance was not informative in this context, for left atrium, right atrium, and right ventricle we retained the first two PCs. To ensure a comparable degree of variance captured across chambers, we retained the first six PCs for the left ventricle, which explained 97% of the variance, and the first three for the aorta, which explained 96% of the variance (**Table S22)**.

**Excluding PITX2 from AF PRS**

We found that left ventricular myocardial wall thickness was negatively associated with AF PRSs and positively associated with incident AF; this latter finding aligned with prior study^16^. To investigate whether this apparent directional inconsistency was driven by the strong association signal at the 4q25 locus, particularly near PITX2, we performed a sensitivity analysis excluding variants in this region from our AF PRS.

Specifically, we excluded from our AF PRS SNPs located within the PITX2 gene (Chromosome 4: 110,617,423-110,642,123), as well as those located 170 kb upstream. Indeed, several important AF-associated variants, including rs2200733^17^, lie outside the gene itself but near it . The final excluded region therefore spanned chromosome 4: 110,617,423-110,812,123.

**Supplementary Results**

**Sensitivity analyses**

Across all diseases, the reported associations between the PRSs and cardiac IDPs remained generally robust after excluding both prevalent and incident cases, adjusting for established risk factors, and using alternative PRSs for the same diseases **(Supplementary Figure S3, Table S11, S15-S16)**. However, the association between the HF PRS and right atrial and right ventricular volumes changed direction when we further adjusted for CVDs risk factors. The association between AF PRS and cardiac IDPs were robust to the exclusion of PITX2 from AF PRS (**Table S23**).

Considering the association between cardiac IDPs and incident diseases, when excluding events that occurred within one year of the imaging visit, HR estimates were broadly robust **(Supplementary Figure S4, Table S17).** However, CIs widened and some associations (particularly abdominal aortic aneurysm and ischaemic stroke) no longer met significance thresholds, consistent with reduced precision after event-count reduction **(Supplementary Figure S4, Table S17).** Indeed, the number of cases decreased from 959 to 876 for coronary artery disease, from 333 to 300 for atrial fibrillation, from 310 to 296 for heart failure, from 233 to 204 for ischaemic stroke and from 52 to 37 for abdominal aortic aneurysm. When assessing the models that violated the proportional hazards (PH) assumption, the inclusion of a time-varying effect (covariate×log(time)) did not reverse the direction of the associations. However, it indicated that these associations attenuated over time **(Table S24).**

We performed several sensitivity analyses for mediation results. These included: treating BMI, current high cholesterol, current high blood pressure, and current diabetes as post-exposure confounders; using alternative PRSs for the same diseases; defining the control and treatment values using two points from the upper tail of the PRS distribution (3 vs 4); defining them using two points from the lower tail of the distribution (-4 vs -3); and allowing for exposure-mediator interaction. Across these analyses, the indirect effect estimates from the single-mediator analyses were robust, although in some cases statistical significance was lost, particularly when post-exposure confounders were included, likely reflecting the greater complexity of the models (**Supplementary Figure S5, Tables S13, S18-S21**).

In the multiple-mediator analyses, indirect effect estimates remained broadly robust, whereas the proportions mediated were less stable, likely because the total effect lost statistical significance in certain models (**Supplementary Figure S6, Table S14**). As model complexity increased (including accounting for post-exposure confounders and for exposure-mediator interaction), the total effect became less stable (**Table S14**). However, our study focused primarily on indirect effects, which presented robust point estimates despite wider confidence intervals when accounting for post-exposure confounding (**Supplementary Figure S6, Table S14**). This resulted in a loss of statistical significance for heart failure (**Supplementary Figure S6, Table S14**).

When implementing single-mediator analysis adjusting for the other cardiac chambers, we performed two different sensitivity analyses. First, we repeated the analyses using alternative PRSs for the same diseases. Second, to reduce potential overadjustment or collider bias, we adjusted our single-mediator models only for established chambers, both as post-exposure confounders and as baseline covariates: left ventricle for heart failure and coronary artery disease, and left atrium for atrial fibrillation and ischaemic stroke. Estimates of indirect effects were largely consistent; however, in some cases significance was lost, also in this case likely because of greater complexity of the model and the low number of cases (**Tables S25-S28**).
